# Polygenic Risk for Schizophrenia, Brain Structure and Environmental Risk in UK Biobank

**DOI:** 10.1101/2021.04.15.21255587

**Authors:** Xingxing Zhu, Joey Ward, Breda Cullen, Donald M. Lyall, Rona J. Strawbridge, Laura M. Lyall, Daniel J. Smith

## Abstract

Schizophrenia is a heritable neurodevelopmental disorder characterized by neuroanatomical changes in the brain but exactly how increased genetic burden for schizophrenia influences brain structure is unknown. Similarly, the impact of environmental risk factors for schizophrenia on brain structure is not fully understood. We investigated how genetic burden for schizophrenia (indexed by a polygenic risk score, PRS-SCZ) was associated with cortical thickness (CT), cortical surface area (SA), cortical volume (CV) and multiple subcortical structures within 18,147 White British ancestry participants from UK Biobank. We also explored whether environmental risk factors for schizophrenia (cannabis use, childhood trauma, low birth weight and Townsend social deprivation index) exacerbated the impact of PRS-SCZ on brain structure. We found that PRS-SCZ was significantly associated with lower CT in the frontal lobe, insula lobe, lateral orbitofrontal cortex, medial orbitofrontal cortex, posterior cingulate cortex and inferior frontal cortex, as well as reduced SA and CV in the supramarginal cortex and superior temporal cortex, but not with differences in subcortical volumes. When models included environmental risk factors as covariates, PRS-SCZ was only associated with lower SA/CV within the supramarginal cortex, superior temporal cortex and inferior frontal cortex. Moreover, no interactions were observed between PRS-SCZ and each of the environmental risk factors on brain structure. Overall, we identified brain structural correlates of PRS-SCZ predominantly within frontal and temporal regions. Some of these associations were independent of environmental risk factors, suggesting that they may represent biomarkers of genetic risk for schizophrenia.

## Introduction

Schizophrenia is a debilitating and complex psychiatric disorder, affecting about 1% of the population ^1^. It is a heritable condition that arises via a complex interaction of genetic and environmental risk factors ^2^. Neuroimaging studies have reported brain structural alterations in cortical and subcortical regions in people with schizophrenia relative to healthy individuals ^3-6^. These brain changes could be driven by genetic factors and may mediate the effect of genetic risk on the phenotype of schizophrenia ^7-9^. Genome-wide association studies (GWAS) have identified hundreds of significant loci and indicate a polygenic architecture of schizophrenia ^8,10,11^. A polygenic risk score for schizophrenia, which captures the cumulative effect of significant risk loci across the whole genome, is useful in exploring the neurobiology of schizophrenia ^12-14^. To date, little is known about how polygenic risk for schizophrenia influences brain structure, or how environmental risk factors may influence these associations.

Convergent evidence has demonstrated widespread grey matter reductions in individuals with schizophrenia, including lower cortical volume (CV), reduced cortical thickness (CT) and smaller surface area (SA) and subcortical volumes, predominantly in the frontal and temporal regions ^3,15-17^. Longitudinal neuroimaging studies have demonstrated progressive grey matter changes related to schizophrenia ^18-20^ as well as morphological differences that may be present prior to the onset of symptoms ^20,21^. High-risk familial studies have observed brain structural alterations in unaffected relatives of patients with schizophrenia relative to controls, suggesting a genetic contribution ^22-24^. The association between polygenic risk scores for schizophrenia (PRS-SCZ) and brain structure has been the subject of some early investigations. Initial work focused on total brain volume, total grey/white matter volume, lateral ventricular volume and subcortical volumes ^25,26^, and a systematic review of seven studies found inconsistent findings plus a lack of significant associations ^27^. The biggest study (N = 14,701) to date reported significant negative associations between genetic risk for schizophrenia and the right pallidum and right thalamus volumes^13^. Studies on cortical measures have also reported negative associations between PRS-SCZ and global CT and insular lobe CT ^12^, frontal and temporal cortex CT^5^, rostral anterior cingulate cortex CT ^28^ within the general population, as well as global cortical thinning in patients with schizophrenia and bipolar disorder ^29^. Relatively few studies have systematically examined associations between PRS-SCZ and regional CT, SA and CV ^5,12,28^. In addition, none of published studies in this area have taken account of the possible influence of environmental risk factors for schizophrenia on brain structure ^5,12,28^.

Environmental risk factors, along with their interaction with genetic risk, contribute to the occurrence and development of schizophrenia ^2,30,31^. Epidemiological studies have identified that childhood adverse events, cannabis use, obstetric complications, and low socioeconomic status are among the strongest non-genetic risk factors for schizophrenia ^32-36^. These factors may contribute, at least in part, to some of the brain structural abnormalities observed in schizophrenia ^29,37,38^ and may also contribute to gene-environment interactions ^38,39^.

Here we investigate associations between PRS-SCZ and cortical metrics (CV, SA, and CT), as well as subcortical volumes, using data from ∼18,000 White British ancestry individuals from UK Biobank. We also assess the impact of environmental risk factors (childhood trauma, cannabis use, birth weight, and Townsend social deprivation index) on any observed associations between PRS-SCZ and these environmental risk factors. We hypothesized that a higher PRS-SCZ would be associated with lower global CT, thinner frontal and temporal cortices, thinner insula lobe, smaller thalamus and pallidum ^5,12,13^ and more localized brain volume reductions. In addition, we expected that environmental risk factors would influence the association between PRS-SCZ and grey matter atrophy we observed.

## Methods

### Participants

This study was conducted using participants from UK Biobank under approval from the NHS National Research Ethics Service (UK Biobank approved applications #6553 and #17689). Freesurfer data (released by Jan 2020) based on the Desikan-Killiany cortical atlas ^40^ were available for a total of 21,915 participants. Participants were excluded for the following reasons: having a developmental or neurological disorder (See Table S1 for detailed participant exclusion criteria); having withdrawn from the projects; non-White British ancestry base on self-report and genetic data; intracranial volume beyond three standard deviations from the sample mean; and genetic data failing to pass quality control. A total of 18,147 participants were included in this study.

### Derivation of the polygenic risk score for schizophrenia

LDpred was utilized to calculate the PRS-SCZ ^41^, based on the summary statistics from a GWAS specific to schizophrenia ^10^, which included 33,426 schizophrenia patients and 54,065 controls, excluding individuals from UK Biobank. Participants were excluded if over 10% of genetic data was missing; if self-reported sex did not match genetic sex; if purported sex chromosome aneuploidy was reported; and if heterozygosity value was a clear outlier.

### Brain imaging variables

All neuroimaging data were acquired, pre-processed, quality controlled and made available by UK Biobank (https://biobank.ctsu.ox.ac.uk/crystal/crystal/docs/brain_mri.pdf). Details on the acquisition parameters and the imaging protocol are documented online and are described within the protocol paper ^42^. Derived phenotypes were used in this study. The main neuroimaging data consisted of global, lobar and regional values for CT, SA and CV of 33 regions (data of temporal pole are not available) in each hemisphere from the Desikan-Killiany cortical atlas ^40^. Global and lobar values were calculated as per Neilson et al. ^12^. In addition, we analysed subcortical volumes including thalamus, caudate, putamen, pallidum, hippocampus, amygdala, accumbens processed by subcortical volumetric segmentation in Freesurfer. A more detailed description of these variables is provided within supplementary materials.

### Environmental risk factors

Childhood trauma was calculated by summing five online questions on previous adversity (data field 20487 to 20491). These questions cover the self-reported frequency of feeling loved, being physically abused, feeling hated, being sexually molested and being taken to the doctor when needed as a child. Participants answered these questions by selecting “Prefer not to answer”, “Never true”, “Rarely true”, “Sometimes true”, “Often” or “Very often true” (supplementary materials).

Cannabis use (https://biobank.ctsu.ox.ac.uk/crystal/field.cgi?id=20453) was reported via an online question, “Have you taken CANNABIS (marijuana, grass, hash, ganja, blow, draw, skunk, weed, spliff, dope), even if it was a long time ago?”. Participants chose from “Prefer not to answer”, “No”, “Yes, 1-2 times”, “Yes, 3-10 times”, “Yes, 11-100 times” and “Yes, more than 100 times”. We grouped participants into two categories: never (participants who answered “No”) and ever (participants who answered “Yes”, having used cannabis at least once).

Birth weight (https://biobank.ctsu.ox.ac.uk/crystal/field.cgi?id=20022) was reported by participants during a verbal interview. To maximise the number of participants included in analyses, missing values at the initial assessment visit were replaced by those at the first repeat assessment visit or the imaging visit.

The Townsend deprivation index (https://biobank.ctsu.ox.ac.uk/crystal/field.cgi?id=189) is based on postcode reported at baseline assessment in UK Biobank. A greater Townsend index score implies a higher level of neighbourhood social and economic deprivation.

### Statistical analysis

All analyses were carried out using Stata/MP 15.0. For associations between PRS-SCZ and brain structure, PRS-SCZ was set as the independent variable; each brain structural measure was set as an outcome. PRS-SCZ × hemisphere interactions were examined in a repeated measures format to determine whether analysis of left and right homologous structures separately was required, with sex, age, age^2^, hemisphere, ICV, scanner positions on the x, y and z axes, genotype array and the first fifteen genetic principal components included as covariates. For brain measures that did not show PRS-SCZ × hemisphere interaction, we repeated the linear mixed model with hemisphere as a fixed effect and included the same covariates as in analyses testing for interaction effects. If there was a significant interaction, analyses on both lateralised structures would be conducted separately. PRS-SCZ and each brain outcome were rescaled into zero mean and unitary standard deviation. Participants with PRS and each brain outcome beyond three standard deviations were excluded. False Discovery Rate (FDR) correction ^43^, with a significance threshold of p□<□0.05, was applied for each metric individually, by correcting over all eight possible lobar structures, twenty-six parcellations or seven subcortical volumes, using ‘p.adjust’ function in R.

Furthermore, considering the potential impact of environmental risk factors, we investigated whether associations between PRS-SCZ and brain measures were significant when childhood trauma, cannabis use, birth weight and Townsend deprivation index were included as additional covariates. In total, 8,707 individuals had complete data for all four environmental factors. Because the subsample was much smaller compared to that in the main analyses, we wished to clarify whether the reduction in sample size influenced the results. Therefore, we repeated previous analyses in this subsample (N = 8,707) without controlling for environmental risk factors. For further examination of the influence of environmental risk factors on the relationship between PRS-SCZ and brain structure, we excluded 21 individuals who definitely had used antipsychotic medication (self-reported at the imaging visit), considering that antipsychotic medication may also influence brain structure ^44,45^.

We also examined the interaction between PRS-SCZ and each of the environmental risk factors on brain structure. The interaction term, together with main effects of the PRS-SCZ and the environmental risk factor were included in the model, as well as same covariates in the analysis for PRS-SCZ and brain structure. Multiple comparisons correction for interaction effects was done using FDR correction (p□<□0.05) in the same way.

## Results

### Demographics

A total of 18,147 participants (ages 45-80 years, 8,651 males) were included in the analysis. Characteristics of the participants are provided in Table 1. There was no significant sex difference in PRS-SCZ (female = 0.006, male = −0.007, t = 0.942, p = 0.346), but the differences in birth weight (female = 3.278 kg, male = 3.462 kg, t = −16.486, p < 0.001), childhood trauma (female = 1.796, male = 1.544, t = 6.201, p < 0.001) and cannabis use (17.766% females vs 23.902% males, Chi squared 76.093, p < 0.001) were significant.

**Table 1.** Descriptive statistics for demographic variables (N=18,147).

| Measure | N (%) or mean ± SD (range) |
| --- | --- |
| Gender | Male: 8,651 (47.67%) |
| Age (years) | 62.78 ± 7.46 (45 - 80) |
| Townsend deprivation index at recruitment (N = 18130) | -2.11 ± 2.57 (-6.26 - 9.16) |
| Birth weight (Kg; N = 11960) | 3.36 ± 0.61 (0.74 – 6.78) |
| Childhood trauma (N = 13044) | 7.69 ± 1.42 (0 – 18) |
| Antipsychotic medication | No: 17,194 (94.75%)<br>Yes: 38 (0.21%)<br>Missing: 915 (5.04%) |
| Cannabis use | No: 10,556 (58.17%)<br>Yes: 2,741 (15.10%)<br>Prefer not to answer: 20 (0.11%)<br>Missing: 4,830 (26.62%) |

### Associations between PRS-SCZ and cortical thickness

We did not find a significant interaction between PRS-SCZ and hemisphere for either global or regional CT (Table S2) and there was no significant association between PRS-SCZ and global CT (β□=□-0.012, p_corrected_□= 0.160; Table 2 shows significant associations only; Table S3 lists all the results). In the eight lobar structures, CT in the frontal lobe (β□=□-0.018, p_corrected_□= 0.040; Table 2) and insula lobe (β□=□-0.016, p_corrected_□= 0.040) were negatively associated with PRS-SCZ. Moreover, we found significant negative associations between PRS-SCZ and CT in the lateral orbitofrontal cortex (β□=□-0.025, p_corrected_□< 0.001), the medial orbitofrontal cortex (β□=□-0.019, p_corrected_□= 0.013), the posterior cingulate cortex (β□=□-0.021, p_corrected_□= 0.009) and the inferior frontal cortex (β□=□-0.026, p_corrected_□< 0.001). Figure 1A illustrates the associations between PRS-SCZ and regional CT.

**Table 2.** The association between polygenic risk score for schizophrenia and brain structures.

| **Brain structure** | **N** | **$\beta$** | **SE** | **z** | **p<sub>uncorrected</sub>** | **p<sub>corrected</sub>** |
| --- | --- | --- | --- | --- | --- | --- |
| ***Lobes*** |  |  |  |  |  |  |
| ***Cortical thickness*** |  |  |  |  |  |  |
| Frontal lobe | 18,036 | -0.018 | 0.007 | -2.760 | 0.006 | 0.040 |
| Insula lobe | 18,086 | -0.016 | 0.006 | -2.590 | 0.010 | 0.040 |
| ***Parcellations*** |  |  |  |  |  |  |
| ***Cortical thickness*** |  |  |  |  |  |  |
| Lateral orbitofrontal | 18,072 | -0.025 | 0.007 | -3.830 | <0.001 | <0.001 |
| Medial orbitofrontal | 18,074 | -0.019 | 0.006 | -3.030 | 0.002 | 0.028 |
| Posterior cingulate | 18,085 | -0.021 | 0.006 | -3.460 | 0.001 | 0.022 |
| Inferior frontal | 18,038 | -0.026 | 0.006 | -4.100 | <0.001 | <0.001 |
| ***Surface area*** |  |  |  |  |  |  |
| Supramarginal | 18,073 | -0.016 | 0.005 | -3.150 | 0.002 | 0.028 |
| Superior temporal | 18,074 | -0.015 | 0.005 | -3.140 | 0.002 | 0.028 |
| ***Cortical volume*** |  |  |  |  |  |  |
| Supramarginal | 18,074 | -0.016 | 0.005 | -3.190 | 0.001 | 0.022 |
| Superior temporal | 18,062 | -0.018 | 0.005 | -3.580 | <0.001 | <0.001 |

**Figure 1:**
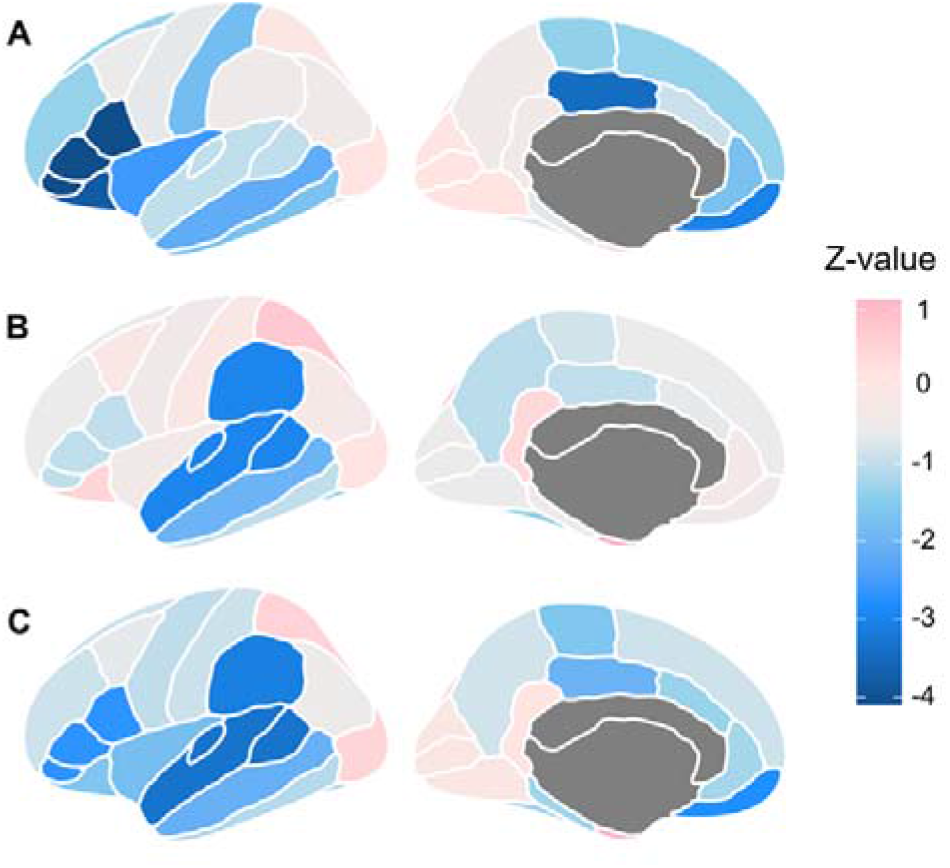
Cortical map of associations between the polygenic risk score for schizophrenia and (A) cortical thickness, (B) surface area and (C) cortical volume. Regions were mapped on the left hemisphere.

In the subsample with complete data on environmental risk factors (childhood trauma, cannabis use, birth weight and Townsend deprivation index) we found modest negative associations between PRS-SCZ and CT in the above regions (Table S4), although none of these associations were significant after FDR correction in this smaller sample. Similarly, after controlling for the environmental risk factors and excluding participants who had used antipsychotic medication, there were no significant associations, although a similar negative pattern of association was found (Table 3 shows significant associations only; Table S5 lists all the results; Figure S1).

**Table 3.** The association between polygenic risk score for schizophrenia and brain structures, adjusted for birth weight, Townsend deprivation index, cannabis use and childhood trauma and excluding participants having used antipsychotics.

| **Brain structure** | **N** | **$\beta$** | **SE** | **z** | **p<sub>uncorrected</sub>** | **p<sub>corrected</sub>** |
| --- | --- | --- | --- | --- | --- | --- |
| ***Parcellations*** |  |  |  |  |  |  |
| ***Surface area*** |  |  |  |  |  |  |
| Supramarginal | 8,638 | -0.028 | 0.007 | -3.910 | <0.001 | <0.001 |
| Superior temporal | 8,637 | -0.021 | 0.007 | -3.090 | 0.002 | 0.026 |
| Inferior frontal | 8,639 | -0.022 | 0.008 | -2.880 | 0.004 | 0.035 |
| ***Cortical volume*** |  |  |  |  |  |  |
| Inferior frontal | 8,635 | -0.026 | 0.008 | -3.330 | 0.001 | 0.026 |

### Associations between PRS-SCZ and surface area

No interactions were observed between PRS-SCZ and hemisphere for global or regional SA (Table S2). Although PRS-SCZ was not associated with global SA (β□=□-0.002, p_corrected_□= 0.672; Table S3; Figure 1B), we found an association with reduced SA in the supramarginal cortex (β□=□-0.016, p_corrected_□= 0.026; Table 2) and the superior temporal cortex (β□=□-0.015, p_corrected_□= 0.026). Analyses in the subsample also found significant associations with these two regions, in addition to inferior frontal cortex (Table S4).

When these analyses were adjusted for environmental factors, the associations with supramarginal cortex (β□=□-0.028, p_corrected_□< 0.001; Table 3), superior temporal cortex (β□=□-0.021, p_corrected_□= 0.026) and inferior frontal cortex (β□=□-0.022, p_corrected_□= 0.035) remained significant.

### Associations between PRS-SCZ and cortical volume

Analyses for the interaction between PRS-SCZ and hemisphere on global or regional CV did not demonstrate any significant effects (Table S2). Similar to the results for surface area, PRS-SCZ was not associated with global CV (β□=□-0.004, p_corrected_□= 0.449; Table S3; Figure 1C), but there were significant negative associations with supramarginal cortex (β□=□-0.016, p_corrected_ = 0.013; Table 2) and superior temporal cortex (β□=□-0.018, p_corrected_□< 0.001).

Analyses in the subsample with complete environmental risk factor data found significant negative associations with the supramarginal cortex and inferior frontal cortex (Table S4). Among them, inferior frontal cortex (β□=□-0.026, p_corrected_□= 0.026; Table 3) remained significant when environmental risk factors were added as covariates.

### Associations between PRS-SCZ and subcortical volumes

Analyses for the interaction between PRS-SCZ and hemisphere found a significant interaction effect for the hippocampus (β□=□0.011, p_corrected_ = 0.025; Table S2) and nucleus accumbens (β□=□-0.015, p_corrected_□= 0.025). Therefore, we also examined the left and right hippocampus and nucleus accumbens separately in following analyses.

PRS-SCZ had a negative association with volume in the right accumbens before (β□=□-0.013, p_uncorrected_□= 0.043; Table S3), but not after FDR correction. Similarly, when environmental risk factors were taken into account, no subcortical structure survived correction for multiple comparisons correction (Table S5).

### PRS-SCZ and environmental risk factors

We firstly tested gene by environment correlation using the linear regression or logistic regression model (cannabis use) for each of the environmental factors. We found PRS-SCZ was significantly associated with Townsend deprivation index (β□=□0.039, t = 1.98, p□= 0.048), cannabis use (odds ratio = 1.077, z = 3.22, p□= 0.001) and childhood trauma (β□=□0.139, t = 6.68, p□< 0.001), but not birth weight (β□=□-0.004, t = −0.62, p□= 0.535).

For the interaction between PRS-SCZ and environmental risk factors, we found no statistically significant results, albeit some modest associations before FDR correction. For example, PRS-SCZ×childhood trauma on SA in the medial orbitofrontal cortex (β□=□-0.006, p_uncorrected_□= 0.013; Table S6): there was a negative association of PRS-SCZ with SA in the medial orbitofrontal cortex in participants exposed to more childhood trauma and a positive association in those who experienced less childhood trauma. More detailed results for the interaction effect are shown in Table S6-S9. Statistics for main effects of the environmental factors are also given (table S10-S13).

## Discussion

In this study we have observed associations between an increased genetic liability for developing schizophrenia and wide-spread cortical differences in generally population, including lower CT in frontal lobe, insula lobe, lateral orbitofrontal cortex, medial orbitofrontal cortex, inferior frontal cortex and posterior cingulate cortex, as well as reduced SA and CV in the supramarginal cortex and superior temporal cortex. In addition, the associations with reduced SA and CV remained after adjustment for environmental risk factors, indicating that some cortical alterations might be driven predominantly by genetic liability for schizophrenia rather than environmental risk factors. Finally, our results suggest that PRS-SCZ and these environmental risk factors may contribute independently to brain abnormalities in schizophrenia. Overall, our findings have expanded on previous findings by making use of data within a much larger population-based sample.

### PRS-SCZ and CT

Previous studies of the relationship between PRS-SCZ and global CT have been inconsistent, with global cortical thinning ^12,29^ or no association ^46^ reported. Here, we found no association between PRS-SCZ and global CT, which is unsurprising because schizophrenia-associated genetic variants showed no significant enrichment in mean CT ^9^. For regional measures, we replicated significant associations with reduced CT in insula, lateral orbitofrontal cortex and inferior frontal gyrus, as reported in previous studies in UK Biobank ^5,12,47^. In addition, we observed reduced CT within the frontal lobe, medial orbitofrontal cortex and posterior cingulate cortex. Reduced CT in these regions has been commonly observed in individuals with schizophrenia ^3,17,48-50^ and in individuals at high genetic risk of schizophrenia ^24,48,51^. These regions may also be implicated in the symptoms seen in schizophrenia: the insula is engaged in auditory hallucinations ^47^; the orbitofrontal cortex is thought to play a role in negative symptoms ^52^; and the frontal lobe has shown associations with cognitive impairment in schizophrenia ^53^.

### PRS-SCZ and SA

We found that PRS-SCZ was not associated with global SA. Regarding this, previous studies have reported inconsistent findings^12,46,54^. Analyses for regional SA found negative associations within temporal cortices: the supramarginal cortex and superior temporal cortex. This is in line with the significant enrichment of schizophrenia GWAS loci in SA of temporal regions ^9^. Previous studies support structural differences in superior temporal gyrus and inferior frontal cortex as possible biomarkers in schizophrenia ^55,56^. Baseline grey matter reductions in superior temporal and inferior frontal areas are associated with later transition to psychosis ^21^. Additionally, correlations have been observed between superior temporal gyrus atrophy and positive psychotic symptoms, especially auditory verbal hallucinations ^57-59^. Similarly, the involvement of supramarginal and inferior frontal cortex in auditory hallucinations has been extensively documented ^60^ and the supramarginal gyrus is associated with delusions of reference and persecutory delusions ^47^. Alterations in these regions may represent general susceptibility to psychotic symptoms and vulnerability to developing schizophrenia.

### PRS-SCZ and CV

For the relationship between PRS-SCZ and brain volume, findings were also contradictory. Van Scheltinga et al. ^25^ found PRS-SCZ was significantly associated with smaller total brain volume regardless of disease status, but later studies found different results even within larger samples ^12,26,61,62^. We found no such association. However, we found significant volume reduction in supramarginal cortex and superior temporal cortex. When environmental risk factors were taken into account, an association with inferior frontal cortex volume was significant, and both supramarginal cortex and superior temporal cortex were nominally significant. As indicated earlier, this is in line with putative roles for these structures in processing auditory inputs, and their relation to auditory hallucinations in schizophrenia ^57,63,64^.

### PRS-SCZ and subcortical volumes

For subcortical volumes, although some studies have reported associations of PRS-SCZ with smaller thalamus and pallidum volume ^13,26,54,65^, others have found no association ^5,46,66^. Here, we observed no association with thalamus or pallidum volumes and a modest association with the right nucleus accumbens. Smaller nucleus accumbens has been reported in individuals with schizophrenia ^4,5^, as well as in their first-degree relatives ^22,48^. The nucleus accumbens is implicated in numerous neurological and psychiatric disorders, and is a main target of antipsychotic drugs and neurosurgical intervention in schizophrenia ^67,68^. However, it should be noted that the association with the right nucleus accumbens was no longer significant after FDR correction. Overall, our data suggest that subcortical brain volumes are not strongly associated with genetic mechanisms of schizophrenia.

### Influence of environmental risk factors

When environmental risk factors were included in the model as covariates and participants on antipsychotic medication were excluded, we observed a similar pattern of grey matter reduction; the effects for the supramarginal cortex, superior temporal cortex and inferior frontal cortex remained significant. Although these environmental risk factors seem to show little impact, they are still important considerations. Marsman et al. ^69^ examined the relative contributions of genes and environment to mental health and found that familial and environmental factors explained around 17% of the variance, of which around 3% by PRS. Indeed, environmental risk factors, such as cannabis use, childhood trauma and socioeconomic status, increase the risk for multidimensional symptoms in schizophrenia, including cognitive, affective, negative and predominantly positive symptoms such as hallucinations and delusions ^70-73^ and elevate the risk of developing schizophrenia ^74,75^. Moreover, these environmental risk factors exhibit associations with structural differences in widespread brain regions across frontal, parietal, temporal and occipital cortex, as well as in subcortical volumes ^39,76-78^. Our results also showed significant associations between birth weight and the brain measures that were significant in analyses including environmental risk factors as covariates, although the other three environmental factors did not show such associations. Future studies on brain abnormalities in schizophrenia should consider the role of environmental risks more closely.

### Correlation between PRS-SCZ and environmental risk factors

We found that genetic risk for schizophrenia was associated with greater likelihood of using cannabis ^79,80^, experiencing more childhood trauma ^81,82^ and higher socioeconomic deprivation, but not birth weight ^12^. Previous studies on the genetic influences on environmental measures demonstrate considerable heritability ^83^ and GWASs have found many genetic variants associated with Townsend deprivation index ^84,85^, birth weight ^86^ and cannabis use ^87,88^. Other studies also report genetic correlations between Townsend deprivation index, cannabis use and schizophrenia ^80,84,87^. As a result, the associations we observed may be due to shared genetic aetiologies.

### Interaction between PRS-SCZ and environmental risk factors

To our knowledge, only one study so far has reported significant interaction between PRS-SCZ and environmental risk on brain structure: PRS-SCZ was negatively associated with mean CT in male cannabis users, while there was no significant association in male nonusers ^38^. By contrast, studies focused on schizophrenia susceptibility genes especially COMT (catechol-O-methyltransferase) and BDNF (brain-derived neurotrophic factor) demonstrated interactions with obstetric complications, early life stress and cannabis abuse ^89,90^, supporting the role of gene–environment interaction. Together with the modest interaction in this study, while preliminary, we recommend that further confirmatory studies in even larger clinical and non-clinical samples should be conducted to improve understanding of gene-environment interactions in schizophrenia.

### Strengths and Limitations

This study represents the largest neuroimaging PRS-SCZ study to date, identifying associations with multiple brain measures. Nevertheless, it is unclear to what extent these structural differences are causes, consequences or confounders of schizophrenia. Importantly, we also considered the influence of environmental risk factors for schizophrenia and explored gene-by-environment interactions on structural brain measures. Considering the modest significant associations, further studies which take these variables into account will need to be undertaken with larger samples. Furthermore, our study was conducted in White British participants aged from 45 to 80, which may restrict generalization of our findings to the other populations with different ancestry or age range. Finally, the environmental factors in this study were limited to childhood trauma, cannabis use, Townsend deprivation index and birth weight. To develop a fuller picture of gene-by-environment interactions, additional studies on other factors such as migration background ^91^ are required.

## Conclusion

In summary, this study suggests that grey matter reductions in multiple regions, predominantly in frontal and temporal regions, are neuroanatomical correlates of increased genetic liability for schizophrenia. Further, environmental risk factors such as childhood trauma, cannabis use, Townsend deprivation index and birth weight, appear to contribute very little to these associations, plus genetic liability for schizophrenia and these environmental factors contribute independently to brain abnormalities in schizophrenia. Our findings are meaningful in terms of identifying neuroimaging-based biomarkers for schizophrenia and elucidating the complex pathophysiology of schizophrenia.

## Data Availability

We used genetic and phenotypic data from the UK Biobank under the application number #6553 and #17689.

https://www.ukbiobank.ac.uk/

## Acknowledgments

XZ acknowledges financial support from China Scholarship Council. RJS is funded by UKRI Innovation-HDR-UK Fellowship (MR/S003061/1). DJS acknowledges support from a Lister Institute Prize Fellowship and an MRC Mental Health Data Pathfinder Award (MC_PC_17217). JW is funded by DJS Lister Institute Prize Fellowship. LML is supported by a Royal College of Physicians of Edinburgh JMAS Sim Fellowship.

