## Supplementary materials for "Polygenic Risk for Schizophrenia, Brain Structure and Environmental Risk in UK Biobank"

### Supplementary information

**Content:**

Supplementary Methods: page2-3

Figures S1: page4

Tables S1-S13: page5-41

Supplementary References

### MRI preprocessing

The T1 images were processed with FreeSurfer (http://surfer.nmr.mgh.harvard.edu/). The technical details are described in previous publications ^1,2^. Briefly, the processing pipeline includes motion correction and averaging, removal of non-brain tissue, automated Talairach transformation, intensity normalisation, white matter segmentation, cortical surface reconstruction and parcellation. Cortical thickness was computed in FreeSurfer by calculating the closest distance from the gray/white matter boundary and the gray/CSF boundary at each vertex on the tessellated surface. Surface area was calculated by summing the area of the vertices in each region. Cortical volume was calculated as the product of the cortical thickness and surface area.

Considering that previous studies conducted statistics based on the Desikan-Killiany atlas ^3^, and to compare results in this study and those in previous ones, we utilized the Desikan-Killiany atlas, with each hemisphere being parcellated into 34 regions. Furthermore, as Neilson et al. ^4^ have done, we calculated global and lobar values for cortical thickness, surface area and cortical volume: Superior Temporal Gyrus (STG) = banks of the superior temporal sulcus, transverse temporal, superior temporal; Inferior Frontal Gyrus (IFG) = pars opercularis, pars triangularis, pars orbitalis; Dorsal Lateral Prefrontal Cortex (DLPFC) = rostral middle frontal, superior frontal; Medial Occipital (MO) = pericalcarine, cuneus, lingual. Similarly, Frontal lobe –DLPFC, Caudal middle frontal gyrus, IFG, Lateral and Medial orbitofrontal cortex, Frontal pole, Precentral gyrus; Temporal lobe – Entorhinal cortex, Parahippocampal gyrus, Temporal pole, Fusiform gyrus and STG; Occipital Lobe – MO and Lateral occipital cortex; Parietal lobe – Superior Parietal cortex, Inferior Parietal cortex, Supramarginal gyrus, Precuneus cortex; Cingulate cortex – Rostral anterior cingulate, Caudal anterior cingulate, Posterior Cingulate, Isthmus division; Insular lobe - Insula; Postcentral lobule – Postcentral gyrus; Paracentral lobule – Paracentral lobule.

### Measures of covariates

Genetic principal components 1–15 calculated via principal component analysis were included as population stratification covariates. Head positions in scanner were also corrected for, including lateral (<https://biobank.ctsu.ox.ac.uk/showcase/field.cgi?id=25756>), transverse (<http://biobank.ctsu.ox.ac.uk/crystal/field.cgi?id=25757>) and longitudinal (<https://biobank.ctsu.ox.ac.uk/showcase/field.cgi?id=25758>) co-ordinates of the centre of the brain mask.

The score of childhood traumatic events was calculated via summing five online questions (<http://biobank.ndph.ox.ac.uk/showcase/label.cgi?id=145>): (1) "When I was growing up... I felt loved"; (2) "When I was growing up... People in my family hit me so hard that it left me with bruises or marks"; (3) "When I was growing up... I felt that someone in my family hated me"; (4) "When I was growing up... Someone molested me (sexually)"; (5) "When I was growing up... There was someone to take me to the doctor if I needed it". Participants answered by selecting “Prefer not to answer”, “Never true”, “Rarely true”, “Sometimes true”, “Often” or “Very often true”. The scores for “felling loved” and “being taken to doctor when needed” are in an opposite direction, so they were reversed when calculating a sum score.

#### Figure S1. Cortical map of regions associated with polygenic risk score for schizophrenia.  (A) results for cortical thickness. (B) results for surface area. (C) results for cortical volume. Warm colours reflect positive association and cool colours reflect negative association.

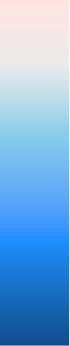

-3

-2

-1

0

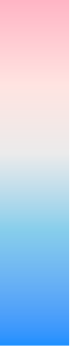

1

**A**

**C**

**B**

Z-value

#### Table S1. Participant exclusion criteria according to self-reported cancer and non-cancer illness (data field 20001 and 20002).

Benign neuroma

Brain abscess/intracranial abscess

Brain cancer/primary malignant brain tumour

Brain haemorrhage

Cerebral aneurysm

Cerebral palsy

Chronic/degenerative neurological problem

dementia/alzheimers/cognitive impairment

Encephalitis

Epilepsy

Fracture skull/head

Head injury

Ischaemic stroke

Meningeal cancer/malignant meningioma

Meningioma/benign meningeal tumour

Meningitis

Motor Neurone Disease

Multiple Sclerosis

Nervous system infection

Neurological injury/trauma

Other demyelinating disease (not Multiple Sclerosis)

Other neurological problem

Parkinson’s Disease

Spina Bifida

Stroke

Subarachnoid haemorrhage

Subdural haemorrhage/haematoma

Transient ischaemic attack

#### Table S2. The interaction between polygenic risk score for schizophrenia and hemisphere on regional cortical thickness, surface area, cortical volume and subcortical volumes.

| **Brain structure** | **β** | **SE** | **z** | **p_uncorrected_** | **p_corrected_** |
| --- | --- | --- | --- | --- | --- |
| ***Global*** |  |  |  |  |  |
| Cortical thickness | 0.008 | 0.005 | 1.570 | 0.116 | 0.174 |
| Surface area | -0.001 | 0.001 | -1.240 | 0.216 | 0.216 |
| Cortical volume | 0.005 | 0.003 | 1.700 | 0.090 | 0.174 |
| ***Lobes*** |  |  |  |  |  |
| ***Cortical thickness*** |  |  |  |  |  |
| Frontal lobe | 0.007 | 0.005 | 1.270 | 0.203 | 0.308 |
| Temporal lobe | 0.008 | 0.007 | 1.270 | 0.203 | 0.308 |
| Occipital lobe | 0.018 | 0.006 | 3.080 | 0.002 | 0.016 |
| Parietal lobe | 0.006 | 0.004 | 1.350 | 0.176 | 0.308 |
| Cingulate lobe | 0.002 | 0.008 | 0.270 | 0.789 | 0.902 |
| Insula lobe | 0.013 | 0.008 | 1.750 | 0.080 | 0.308 |
| Postcentral lobe | <0.001 | 0.006 | -0.080 | 0.937 | 0.937 |
| Paracentral lobe | 0.007 | 0.006 | 1.200 | 0.231 | 0.308 |
| ***Surface area*** |  |  |  |  |  |
| Frontal lobe | -0.004 | 0.002 | -1.740 | 0.082 | 0.328 |
| Temporal lobe | -0.002 | 0.003 | -0.840 | 0.403 | 0.537 |
| Occipital lobe | -0.001 | 0.003 | -0.220 | 0.827 | 0.827 |
| Parietal lobe | 0.005 | 0.003 | 1.460 | 0.143 | 0.381 |
| Cingulate lobe | 0.008 | 0.006 | 1.300 | 0.192 | 0.384 |
| Insula lobe | -0.002 | 0.006 | -0.330 | 0.742 | 0.827 |
| Postcentral lobe | -0.009 | 0.005 | -1.780 | 0.076 | 0.328 |
| Paracentral lobe | -0.006 | 0.006 | -0.940 | 0.347 | 0.537 |
| ***Cortical volume*** |  |  |  |  |  |
| Frontal lobe | <0.001 | 0.003 | 0.140 | 0.888 | 0.922 |
| Temporal lobe | 0.005 | 0.004 | 1.280 | 0.199 | 0.360 |
| Occipital lobe | 0.007 | 0.004 | 1.450 | 0.147 | 0.360 |
| Parietal lobe | 0.008 | 0.004 | 2.170 | 0.030 | 0.240 |
| Cingulate lobe | 0.009 | 0.007 | 1.240 | 0.214 | 0.360 |
| Insula lobe | 0.006 | 0.005 | 1.210 | 0.225 | 0.360 |
| Postcentral lobe | -0.007 | 0.006 | -1.090 | 0.276 | 0.368 |
| Paracentral lobe | -0.001 | 0.007 | -0.100 | 0.922 | 0.922 |
| ***Parcellations*** |  |  |  |  |  |
| ***Cortical thickness*** |  |  |  |  |  |
| Caudal anterior cingulate | 0.008 | 0.009 | 0.900 | 0.368 | 0.563 |
| Caudal middle frontal | 0.012 | 0.006 | 2.050 | 0.041 | 0.323 |
| Entorhinal | 0.001 | 0.008 | 0.170 | 0.866 | 0.937 |
| Fusiform | 0.009 | 0.007 | 1.290 | 0.196 | 0.451 |
| Inferior parietal | 0.008 | 0.006 | 1.380 | 0.168 | 0.451 |
| Inferior temporal | 0.014 | 0.007 | 1.910 | 0.057 | 0.323 |
| Isthmus cingulate | -0.011 | 0.007 | -1.460 | 0.144 | 0.451 |
| Lateral occipital | 0.016 | 0.006 | 2.580 | 0.010 | 0.130 |
| Lateral orbitofrontal | 0.005 | 0.007 | 0.680 | 0.495 | 0.667 |
| Medial orbitofrontal | 0.001 | 0.007 | 0.080 | 0.935 | 0.937 |
| Middle temporal | 0.009 | 0.007 | 1.250 | 0.212 | 0.451 |
| Parahippocampal | 0.004 | 0.007 | 0.530 | 0.598 | 0.707 |
| Paracentral | 0.007 | 0.006 | 1.200 | 0.231 | 0.451 |
| Postcentral | <0.001 | 0.006 | -0.080 | 0.937 | 0.937 |
| Posterior cingulate | -0.004 | 0.008 | -0.560 | 0.576 | 0.707 |
| Precentral | 0.001 | 0.005 | 0.170 | 0.861 | 0.937 |
| Precuneus | 0.004 | 0.005 | 0.830 | 0.404 | 0.584 |
| Rostral anterior cingulate | 0.006 | 0.009 | 0.650 | 0.513 | 0.667 |
| Superior parietal | 0.006 | 0.005 | 1.170 | 0.243 | 0.451 |
| Supramarginal | 0.006 | 0.006 | 0.980 | 0.325 | 0.528 |
| Frontal pole | 0.015 | 0.008 | 1.840 | 0.066 | 0.323 |
| Insula | 0.013 | 0.008 | 1.750 | 0.080 | 0.323 |
| Superior temporal | 0.011 | 0.006 | 1.710 | 0.087 | 0.323 |
| Inferior frontal | 0.008 | 0.006 | 1.350 | 0.179 | 0.451 |
| Dorsal lateral prefrontal | 0.005 | 0.005 | 1.020 | 0.307 | 0.528 |
| Medial occipital | 0.016 | 0.006 | 2.620 | 0.009 | 0.130 |
| ***Surface area*** |  |  |  |  |  |
| Caudal anterior cingulate | 0.013 | 0.009 | 1.470 | 0.143 | 0.530 |
| Caudal middle frontal | -0.003 | 0.006 | -0.400 | 0.687 | 0.968 |
| Entorhinal | 0.014 | 0.007 | 2.140 | 0.032 | 0.329 |
| Fusiform | -0.008 | 0.005 | -1.400 | 0.163 | 0.530 |
| Inferior parietal | -0.002 | 0.006 | -0.260 | 0.792 | 0.968 |
| Inferior temporal | -0.006 | 0.006 | -1.040 | 0.299 | 0.777 |
| Isthmus cingulate | 0.003 | 0.006 | 0.570 | 0.572 | 0.968 |
| Lateral occipital | <0.001 | 0.005 | -0.090 | 0.931 | 0.968 |
| Lateral orbitofrontal | -0.001 | 0.005 | -0.290 | 0.771 | 0.968 |
| Medial orbitofrontal | 0.002 | 0.006 | 0.300 | 0.762 | 0.968 |
| Middle temporal | -0.010 | 0.005 | -2.070 | 0.038 | 0.329 |
| Parahippocampal | -0.008 | 0.007 | -1.180 | 0.238 | 0.688 |
| Paracentral | -0.006 | 0.006 | -0.940 | 0.347 | 0.800 |
| Postcentral | -0.009 | 0.005 | -1.780 | 0.076 | 0.329 |
| Posterior cingulate | 0.015 | 0.007 | 2.130 | 0.033 | 0.329 |
| Precentral | -0.001 | 0.005 | -0.160 | 0.871 | 0.968 |
| Precuneus | <0.001 | 0.004 | -0.100 | 0.919 | 0.968 |
| Rostral anterior cingulate | -0.006 | 0.008 | -0.750 | 0.453 | 0.841 |
| Superior parietal | 0.005 | 0.005 | 0.900 | 0.369 | 0.800 |
| Supramarginal | 0.012 | 0.006 | 1.920 | 0.055 | 0.329 |
| Frontal pole | 0.006 | 0.008 | 0.780 | 0.436 | 0.841 |
| Insula | -0.002 | 0.006 | -0.330 | 0.742 | 0.968 |
| Superior temporal | 0.001 | 0.005 | 0.130 | 0.899 | 0.968 |
| Inferior frontal | 0.001 | 0.006 | 0.140 | 0.890 | 0.968 |
| Dorsal lateral prefrontal | -0.006 | 0.003 | -1.770 | 0.076 | 0.329 |
| Medial occipital | <0.001 | 0.004 | 0.010 | 0.994 | 0.994 |
| ***Cortical volume*** |  |  |  |  |  |
| Caudal anterior cingulate | 0.014 | 0.010 | 1.420 | 0.155 | 0.650 |
| Caudal middle frontal | 0.001 | 0.006 | 0.170 | 0.863 | 0.929 |
| Entorhinal | 0.009 | 0.007 | 1.280 | 0.200 | 0.650 |
| Fusiform | -0.001 | 0.006 | -0.090 | 0.929 | 0.929 |
| Inferior parietal | 0.003 | 0.006 | 0.410 | 0.682 | 0.887 |
| Inferior temporal | -0.001 | 0.006 | -0.230 | 0.816 | 0.929 |
| Isthmus cingulate | 0.004 | 0.007 | 0.600 | 0.551 | 0.799 |
| Lateral occipital | 0.005 | 0.006 | 0.850 | 0.396 | 0.699 |
| Lateral orbitofrontal | 0.002 | 0.005 | 0.520 | 0.602 | 0.824 |
| Medial orbitofrontal | 0.005 | 0.006 | 0.820 | 0.410 | 0.699 |
| Middle temporal | -0.004 | 0.005 | -0.790 | 0.430 | 0.699 |
| Parahippocampal | -0.001 | 0.007 | -0.180 | 0.855 | 0.929 |
| Paracentral | -0.001 | 0.007 | -0.100 | 0.922 | 0.929 |
| Postcentral | -0.007 | 0.006 | -1.090 | 0.276 | 0.671 |
| Posterior cingulate | 0.012 | 0.007 | 1.580 | 0.113 | 0.650 |
| Precentral | -0.001 | 0.005 | -0.110 | 0.913 | 0.929 |
| Precuneus | 0.003 | 0.004 | 0.590 | 0.553 | 0.799 |
| Rostral anterior cingulate | -0.007 | 0.008 | -0.810 | 0.420 | 0.699 |
| Superior parietal | 0.006 | 0.005 | 1.070 | 0.284 | 0.671 |
| Supramarginal | 0.013 | 0.006 | 2.080 | 0.038 | 0.494 |
| Frontal pole | 0.018 | 0.008 | 2.340 | 0.019 | 0.494 |
| Insula | 0.006 | 0.005 | 1.210 | 0.225 | 0.650 |
| Superior temporal | 0.007 | 0.005 | 1.310 | 0.191 | 0.650 |
| Inferior frontal | 0.007 | 0.006 | 1.220 | 0.223 | 0.650 |
| Dorsal lateral prefrontal | -0.004 | 0.004 | -0.990 | 0.323 | 0.699 |
| Medial occipital | 0.008 | 0.005 | 1.580 | 0.115 | 0.650 |
| ***Subcortical volumes*** |  |  |  |  |  |
| Thalamus | 0.001 | 0.003 | 0.430 | 0.664 | 0.695 |
| Caudate | 0.003 | 0.003 | 1.000 | 0.317 | 0.555 |
| Putamen | 0.002 | 0.003 | 0.720 | 0.474 | 0.664 |
| Pallidum | -0.004 | 0.004 | -1.020 | 0.309 | 0.555 |
| Hippocampus | 0.011 | 0.004 | 2.740 | 0.006 | 0.025 |
| Amygdala | -0.002 | 0.005 | -0.390 | 0.695 | 0.695 |
| Accumbens | -0.015 | 0.005 | -2.680 | 0.007 | 0.025 |

#### Table S3. The association between polygenic risk score for schizophrenia and brain structures. The model was conducted with age, age^2^, sex, total ICV, hemisphere, head position coordinates, genotype array and the first fifteen genetic principal components included as covariates.

| **Brain structure** | **N** | **β** | **SE** | **z** | **p** | **p_corrected_** |
| --- | --- | --- | --- | --- | --- | --- |
| ***Global*** |  |  |  |  |  |  |
| Cortical thickness | 18,033 | -0.012 | 0.007 | -1.750 | 0.080 | 0.240 |
| Surface area | 18,054 | -0.002 | 0.004 | -0.420 | 0.672 | 0.672 |
| Cortical volume | 18,067 | -0.004 | 0.004 | -1.040 | 0.299 | 0.449 |
| ***Lobes*** |  |  |  |  |  |  |
| ***Cortical thickness*** |  |  |  |  |  |  |
| Frontal lobe | 18,036 | -0.018 | 0.007 | -2.760 | 0.006 | 0.040 |
| Temporal lobe | 18,061 | -0.008 | 0.006 | -1.220 | 0.221 | 0.295 |
| Occipital lobe | 18,078 | 0.001 | 0.007 | 0.220 | 0.826 | 0.861 |
| Parietal lobe | 18,006 | -0.001 | 0.007 | -0.170 | 0.861 | 0.861 |
| Cingulate lobe | 18,083 | -0.013 | 0.006 | -2.160 | 0.030 | 0.080 |
| Insula lobe | 18,086 | -0.016 | 0.006 | -2.590 | 0.010 | 0.040 |
| Postcentral lobe | 18,076 | -0.012 | 0.007 | -1.850 | 0.065 | 0.130 |
| Paracentral lobe | 18,075 | -0.009 | 0.007 | -1.390 | 0.164 | 0.262 |
| ***Surface area*** |  |  |  |  |  |  |
| Frontal lobe | 18,059 | 0.001 | 0.004 | 0.190 | 0.852 | 0.928 |
| Temporal lobe | 18,064 | -0.008 | 0.004 | -2.040 | 0.041 | 0.328 |
| Occipital lobe | 18,060 | 0.001 | 0.006 | 0.200 | 0.839 | 0.928 |
| Parietal lobe | 18,060 | -0.002 | 0.005 | -0.390 | 0.699 | 0.928 |
| Cingulate lobe | 18,075 | 0.001 | 0.004 | 0.180 | 0.854 | 0.928 |
| Insula lobe | 18,074 | <0.001 | 0.005 | 0.090 | 0.928 | 0.928 |
| Postcentral lobe | 18,074 | 0.002 | 0.005 | 0.410 | 0.684 | 0.928 |
| Paracentral lobe | 18,072 | -0.002 | 0.005 | -0.370 | 0.709 | 0.928 |
| ***Cortical volume*** |  |  |  |  |  |  |
| Frontal lobe | 18,065 | -0.005 | 0.004 | -1.080 | 0.282 | 0.376 |
| Temporal lobe | 18,069 | -0.010 | 0.005 | -2.290 | 0.022 | 0.176 |
| Occipital lobe | 18,070 | 0.006 | 0.006 | 1.090 | 0.276 | 0.376 |
| Parietal lobe | 18,074 | -0.003 | 0.005 | -0.580 | 0.563 | 0.643 |
| Cingulate lobe | 18,077 | -0.006 | 0.005 | -1.210 | 0.226 | 0.376 |
| Insula lobe | 18,054 | -0.008 | 0.005 | -1.460 | 0.143 | 0.376 |
| Postcentral lobe | 18,073 | -0.002 | 0.005 | -0.440 | 0.658 | 0.658 |
| Paracentral lobe | 18,079 | -0.007 | 0.005 | -1.270 | 0.205 | 0.376 |
| ***Parcellations*** |  |  |  |  |  |  |
| ***Cortical thickness*** |  |  |  |  |  |  |
| Caudal anterior cingulate | 18,086 | -0.005 | 0.006 | -0.920 | 0.356 | 0.806 |
| Caudal middle frontal | 18,039 | -0.004 | 0.006 | -0.610 | 0.543 | 0.922 |
| Entorhinal | 18,053 | -0.001 | 0.006 | -0.150 | 0.882 | 0.964 |
| Fusiform | 18,071 | -0.004 | 0.006 | -0.690 | 0.489 | 0.905 |
| Inferior parietal | 18,032 | -0.002 | 0.006 | -0.370 | 0.714 | 0.929 |
| Inferior temporal | 18,079 | -0.011 | 0.006 | -1.730 | 0.084 | 0.389 |
| Isthmus cingulate | 18,083 | -0.003 | 0.006 | -0.410 | 0.684 | 0.929 |
| Lateral occipital | 18,075 | 0.001 | 0.007 | 0.170 | 0.864 | 0.964 |
| Lateral orbitofrontal | 18,072 | -0.025 | 0.007 | -3.830 | <0.001 | <0.001 |
| Medial orbitofrontal | 18,074 | -0.019 | 0.006 | -3.030 | 0.002 | 0.028 |
| Middle temporal | 18,083 | -0.014 | 0.006 | -2.190 | 0.029 | 0.196 |
| Parahippocampal | 18,079 | -0.004 | 0.006 | -0.700 | 0.486 | 0.905 |
| Paracentral | 18,075 | -0.009 | 0.007 | -1.390 | 0.164 | 0.569 |
| Postcentral | 18,076 | -0.012 | 0.007 | -1.850 | 0.065 | 0.361 |
| Posterior cingulate | 18,085 | -0.021 | 0.006 | -3.460 | 0.001 | 0.022 |
| Precentral | 18,053 | -0.004 | 0.006 | -0.660 | 0.509 | 0.911 |
| Precuneus | 18,048 | -0.002 | 0.007 | -0.360 | 0.720 | 0.929 |
| Rostral anterior cingulate | 18,082 | -0.010 | 0.006 | -1.750 | 0.080 | 0.386 |
| Superior parietal | 18,034 | <0.001 | 0.007 | -0.040 | 0.969 | 0.971 |
| Supramarginal | 18,032 | -0.003 | 0.006 | -0.460 | 0.644 | 0.929 |
| Frontal pole | 18,075 | -0.009 | 0.006 | -1.530 | 0.126 | 0.511 |
| Insula | 18,086 | -0.016 | 0.006 | -2.590 | 0.010 | 0.093 |
| Superior temporal | 18,058 | -0.007 | 0.007 | -1.010 | 0.311 | 0.767 |
| Inferior frontal | 18,038 | -0.026 | 0.006 | -4.100 | <0.001 | <0.001 |
| Dorsal lateral prefrontal | 18,001 | -0.009 | 0.006 | -1.360 | 0.174 | 0.585 |
| Medial occipital | 18,076 | 0.001 | 0.007 | 0.180 | 0.857 | 0.964 |
| ***Surface area*** |  |  |  |  |  |  |
| Caudal anterior cingulate | 18,078 | -0.001 | 0.005 | -0.180 | 0.858 | 0.964 |
| Caudal middle frontal | 18,068 | 0.002 | 0.005 | 0.320 | 0.747 | 0.953 |
| Entorhinal | 18,082 | 0.010 | 0.006 | 1.600 | 0.109 | 0.465 |
| Fusiform | 18,076 | -0.004 | 0.005 | -0.930 | 0.353 | 0.806 |
| Inferior parietal | 18,079 | 0.001 | 0.005 | 0.240 | 0.811 | 0.964 |
| Inferior temporal | 18,078 | -0.003 | 0.005 | -0.530 | 0.598 | 0.922 |
| Isthmus cingulate | 18,068 | 0.004 | 0.005 | 0.870 | 0.382 | 0.810 |
| Lateral occipital | 18,071 | 0.003 | 0.005 | 0.650 | 0.519 | 0.914 |
| Lateral orbitofrontal | 18,075 | 0.005 | 0.005 | 0.950 | 0.343 | 0.806 |
| Medial orbitofrontal | 18,075 | <0.001 | 0.004 | 0.050 | 0.958 | 0.971 |
| Middle temporal | 18,068 | -0.007 | 0.005 | -1.520 | 0.129 | 0.511 |
| Parahippocampal | 18,077 | <0.001 | 0.005 | 0.040 | 0.965 | 0.971 |
| Paracentral | 18,072 | -0.002 | 0.005 | -0.370 | 0.709 | 0.929 |
| Postcentral | 18,074 | 0.002 | 0.005 | 0.410 | 0.684 | 0.929 |
| Posterior cingulate | 18,070 | -0.003 | 0.005 | -0.530 | 0.593 | 0.922 |
| Precentral | 18,072 | <0.001 | 0.005 | 0.050 | 0.960 | 0.971 |
| Precuneus | 18,057 | -0.003 | 0.005 | -0.580 | 0.563 | 0.922 |
| Rostral anterior cingulate | 18,077 | 0.001 | 0.005 | 0.160 | 0.869 | 0.964 |
| Superior parietal | 18,065 | 0.006 | 0.005 | 1.190 | 0.232 | 0.660 |
| Supramarginal | 18,073 | -0.016 | 0.005 | -3.150 | 0.002 | 0.028 |
| Frontal pole | 18,084 | 0.004 | 0.005 | 0.740 | 0.462 | 0.884 |
| Insula | 18,074 | <0.001 | 0.005 | 0.090 | 0.928 | 0.971 |
| Superior temporal | 18,074 | -0.015 | 0.005 | -3.140 | 0.002 | 0.028 |
| Inferior frontal | 18,067 | -0.003 | 0.005 | -0.530 | 0.596 | 0.922 |
| Dorsal lateral prefrontal | 18,058 | <0.001 | 0.004 | -0.090 | 0.929 | 0.971 |
| Medial occipital | 18,067 | -0.001 | 0.006 | -0.120 | 0.906 | 0.971 |
| ***Cortical volume*** |  |  |  |  |  |  |
| Caudal anterior cingulate | 18,086 | -0.005 | 0.005 | -0.930 | 0.355 | 0.806 |
| Caudal middle frontal | 18,072 | -0.001 | 0.005 | -0.140 | 0.886 | 0.964 |
| Entorhinal | 18,075 | 0.011 | 0.006 | 1.910 | 0.057 | 0.352 |
| Fusiform | 18,079 | -0.004 | 0.005 | -0.800 | 0.423 | 0.824 |
| Inferior parietal | 18,078 | <0.001 | 0.005 | -0.040 | 0.971 | 0.971 |
| Inferior temporal | 18,079 | -0.003 | 0.005 | -0.670 | 0.505 | 0.911 |
| Isthmus cingulate | 18,079 | 0.004 | 0.005 | 0.810 | 0.418 | 0.824 |
| Lateral occipital | 18,075 | 0.006 | 0.005 | 1.150 | 0.251 | 0.697 |
| Lateral orbitofrontal | 18,063 | -0.007 | 0.005 | -1.270 | 0.203 | 0.632 |
| Medial orbitofrontal | 18,081 | -0.013 | 0.005 | -2.570 | 0.010 | 0.093 |
| Middle temporal | 18,073 | -0.009 | 0.005 | -1.790 | 0.073 | 0.386 |
| Parahippocampal | 18,082 | -0.005 | 0.006 | -0.850 | 0.394 | 0.810 |
| Paracentral | 18,079 | -0.007 | 0.005 | -1.270 | 0.205 | 0.632 |
| Postcentral | 18,073 | -0.002 | 0.005 | -0.440 | 0.658 | 0.929 |
| Posterior cingulate | 18,080 | -0.009 | 0.005 | -1.710 | 0.088 | 0.391 |
| Precentral | 18,073 | -0.003 | 0.005 | -0.560 | 0.574 | 0.922 |
| Precuneus | 18,055 | -0.002 | 0.005 | -0.370 | 0.711 | 0.929 |
| Rostral anterior cingulate | 18,084 | -0.004 | 0.005 | -0.820 | 0.411 | 0.824 |
| Superior parietal | 18,078 | 0.007 | 0.006 | 1.220 | 0.223 | 0.660 |
| Supramarginal | 18,074 | -0.016 | 0.005 | -3.190 | 0.001 | 0.022 |
| Frontal pole | 18,061 | -0.002 | 0.006 | -0.300 | 0.767 | 0.964 |
| Insula | 18,054 | -0.008 | 0.005 | -1.460 | 0.143 | 0.529 |
| Superior temporal | 18,062 | -0.018 | 0.005 | -3.580 | <0.001 | <0.001 |
| Inferior frontal | 18,068 | -0.013 | 0.005 | -2.490 | 0.013 | 0.103 |
| Dorsal lateral prefrontal | 18,068 | -0.002 | 0.005 | -0.410 | 0.680 | 0.929 |
| Medial occipital | 18,066 | 0.004 | 0.006 | 0.600 | 0.550 | 0.922 |
| ***Subcortical volumes*** |  |  |  |  |  |  |
| thalamus | 18,054 | -0.004 | 0.005 | -0.860 | 0.392 | 0.719 |
| caudate | 18,026 | -0.001 | 0.006 | -0.150 | 0.882 | 0.882 |
| putamen | 18,029 | -0.001 | 0.006 | -0.240 | 0.810 | 0.882 |
| pallidum | 18,061 | -0.006 | 0.005 | -1.010 | 0.310 | 0.719 |
| hippocampus | 18,064 | -0.003 | 0.006 | -0.560 | 0.576 | 0.882 |
| amygdala | 18,066 | -0.005 | 0.005 | -0.900 | 0.371 | 0.719 |
| accumbens | 18,066 | -0.006 | 0.006 | -1.040 | 0.299 | 0.719 |
| left hippocampus | 18,014 | -0.009 | 0.006 | -1.470 | 0.141 | 0.719 |
| right hippocampus | 18,012 | 0.002 | 0.006 | 0.400 | 0.690 | 0.882 |
| left accumbens | 18,002 | 0.002 | 0.006 | 0.250 | 0.802 | 0.882 |
| right accumbens | 18,005 | -0.013 | 0.006 | -2.030 | 0.043 | 0.473 |

#### Table S4. The association between polygenic risk score for schizophrenia and brain structures in the subsample with complete environmental risk data.

| **Brain structure** | **N** | **β** | **SE** | **t** | **p_uncorrected_** | **p_corrected_** |
| --- | --- | --- | --- | --- | --- | --- |
| ***Global*** |  |  |  |  |  |  |
| Cortical thickness | 8,643 | 0.001 | 0.009 | 0.060 | 0.949 | 0.949 |
| Surface area | 8,654 | -0.010 | 0.005 | -1.820 | 0.068 | 0.204 |
| Cortical volume | 8,658 | -0.005 | 0.006 | -0.840 | 0.399 | 0.599 |
| ***Lobes*** |  |  |  |  |  |  |
| ***Cortical thickness*** |  |  |  |  |  |  |
| Frontal lobe | 8,644 | -0.006 | 0.009 | -0.680 | 0.494 | 0.693 |
| Temporal lobe | 8,657 | 0.001 | 0.009 | 0.150 | 0.879 | 0.962 |
| Occipital lobe | 8,661 | 0.012 | 0.010 | 1.290 | 0.196 | 0.618 |
| Parietal lobe | 8,631 | 0.010 | 0.010 | 1.060 | 0.291 | 0.618 |
| Cingulate lobe | 8,663 | -0.010 | 0.009 | -1.160 | 0.245 | 0.618 |
| Insula lobe | 8,666 | -0.009 | 0.009 | -1.020 | 0.309 | 0.618 |
| Postcentral lobe | 8,664 | 0.006 | 0.009 | 0.640 | 0.520 | 0.693 |
| Paracentral lobe | 8,660 | <0.001 | 0.009 | 0.050 | 0.962 | 0.962 |
| ***Surface area*** |  |  |  |  |  |  |
| Frontal lobe | 8,655 | -0.007 | 0.006 | -1.150 | 0.248 | 0.440 |
| Temporal lobe | 8,655 | -0.016 | 0.006 | -2.640 | 0.008 | 0.064 |
| Occipital lobe | 8,653 | -0.005 | 0.008 | -0.590 | 0.553 | 0.632 |
| Parietal lobe | 8,657 | -0.010 | 0.007 | -1.440 | 0.150 | 0.440 |
| Cingulate lobe | 8,660 | -0.007 | 0.006 | -1.110 | 0.269 | 0.440 |
| Insula lobe | 8,662 | -0.008 | 0.007 | -1.090 | 0.275 | 0.440 |
| Postcentral lobe | 8,660 | -0.003 | 0.007 | -0.470 | 0.638 | 0.638 |
| Paracentral lobe | 8,660 | -0.005 | 0.007 | -0.680 | 0.497 | 0.632 |
| ***Cortical volume*** |  |  |  |  |  |  |
| Frontal lobe | 8,659 | -0.005 | 0.006 | -0.870 | 0.385 | 0.576 |
| Temporal lobe | 8,657 | -0.012 | 0.006 | -1.830 | 0.067 | 0.189 |
| Occipital lobe | 8,659 | 0.006 | 0.008 | 0.790 | 0.432 | 0.576 |
| Parietal lobe | 8,662 | -0.004 | 0.007 | -0.590 | 0.557 | 0.614 |
| Cingulate lobe | 8,660 | -0.013 | 0.007 | -1.900 | 0.057 | 0.189 |
| Insula lobe | 8,650 | -0.013 | 0.007 | -1.810 | 0.071 | 0.189 |
| Postcentral lobe | 8,663 | 0.006 | 0.008 | 0.820 | 0.412 | 0.576 |
| Paracentral lobe | 8,662 | -0.004 | 0.008 | -0.500 | 0.614 | 0.614 |
| ***Parcellations*** |  |  |  |  |  |  |
| ***Cortical thickness*** |  |  |  |  |  |  |
| Caudal anterior cingulate | 8,665 | -0.009 | 0.008 | -1.130 | 0.258 | 0.676 |
| Caudal middle frontal | 8,644 | 0.014 | 0.009 | 1.540 | 0.124 | 0.645 |
| Entorhinal | 8,653 | -0.006 | 0.009 | -0.650 | 0.519 | 0.784 |
| Fusiform | 8,659 | 0.010 | 0.009 | 1.090 | 0.275 | 0.676 |
| Inferior parietal | 8,643 | 0.007 | 0.009 | 0.720 | 0.474 | 0.784 |
| Inferior temporal | 8,663 | -0.004 | 0.009 | -0.430 | 0.670 | 0.830 |
| Isthmus cingulate | 8,663 | -0.001 | 0.009 | -0.140 | 0.889 | 1.000 |
| Lateral occipital | 8,658 | 0.011 | 0.010 | 1.140 | 0.252 | 0.676 |
| Lateral orbitofrontal | 8,659 | -0.022 | 0.009 | -2.420 | 0.016 | 0.399 |
| Medial orbitofrontal | 8,659 | -0.017 | 0.009 | -1.840 | 0.066 | 0.429 |
| Middle temporal | 8,664 | -0.001 | 0.009 | -0.070 | 0.942 | 1.000 |
| Parahippocampal | 8,664 | <0.001 | 0.009 | <0.001 | 1.000 | 1.000 |
| Paracentral | 8,660 | <0.001 | 0.009 | 0.050 | 0.962 | 1.000 |
| Postcentral | 8,664 | 0.006 | 0.009 | 0.640 | 0.520 | 0.784 |
| Posterior cingulate | 8,666 | -0.018 | 0.009 | -2.050 | 0.041 | 0.399 |
| Precentral | 8,652 | 0.005 | 0.009 | 0.560 | 0.573 | 0.784 |
| Precuneus | 8,650 | 0.007 | 0.010 | 0.710 | 0.477 | 0.784 |
| Rostral anterior cingulate | 8,663 | -0.003 | 0.008 | -0.340 | 0.733 | 0.866 |
| Superior parietal | 8,644 | 0.011 | 0.010 | 1.120 | 0.264 | 0.676 |
| Supramarginal | 8,645 | 0.009 | 0.009 | 1.010 | 0.312 | 0.676 |
| Frontal pole | 8,664 | -0.005 | 0.009 | -0.600 | 0.547 | 0.784 |
| Insula | 8,666 | -0.009 | 0.009 | -1.020 | 0.309 | 0.676 |
| Superior temporal | 8,655 | 0.007 | 0.010 | 0.770 | 0.444 | 0.784 |
| Inferior frontal | 8,643 | -0.018 | 0.009 | -1.990 | 0.046 | 0.399 |
| Dorsal lateral prefrontal | 8,634 | 0.004 | 0.009 | 0.480 | 0.633 | 0.823 |
| Medial occipital | 8,660 | 0.010 | 0.010 | 1.050 | 0.293 | 0.676 |
| ***Surface area*** |  |  |  |  |  |  |
| Caudal anterior cingulate | 8,661 | -0.008 | 0.007 | -1.100 | 0.272 | 0.558 |
| Caudal middle frontal | 8,658 | 0.001 | 0.008 | 0.100 | 0.918 | 0.963 |
| Entorhinal | 8,667 | 0.008 | 0.009 | 0.910 | 0.364 | 0.592 |
| Fusiform | 8,659 | -0.008 | 0.007 | -1.170 | 0.244 | 0.558 |
| Inferior parietal | 8,662 | 0.007 | 0.007 | 0.940 | 0.349 | 0.592 |
| Inferior temporal | 8,659 | -0.012 | 0.007 | -1.690 | 0.091 | 0.473 |
| Isthmus cingulate | 8,657 | -0.006 | 0.007 | -0.800 | 0.423 | 0.647 |
| Lateral occipital | 8,657 | 0.002 | 0.008 | 0.220 | 0.827 | 0.935 |
| Lateral orbitofrontal | 8,661 | -0.001 | 0.007 | -0.080 | 0.934 | 0.963 |
| Medial orbitofrontal | 8,661 | -0.004 | 0.006 | -0.570 | 0.572 | 0.744 |
| Middle temporal | 8,653 | -0.012 | 0.007 | -1.770 | 0.077 | 0.473 |
| Parahippocampal | 8,661 | -0.011 | 0.008 | -1.380 | 0.168 | 0.558 |
| Paracentral | 8,660 | -0.005 | 0.007 | -0.680 | 0.497 | 0.718 |
| Postcentral | 8,660 | -0.003 | 0.007 | -0.470 | 0.638 | 0.790 |
| Posterior cingulate | 8,660 | -0.010 | 0.007 | -1.380 | 0.168 | 0.558 |
| Precentral | 8,658 | <0.001 | 0.007 | 0.050 | 0.963 | 0.963 |
| Precuneus | 8,655 | -0.008 | 0.007 | -1.080 | 0.279 | 0.558 |
| Rostral anterior cingulate | 8,660 | -0.004 | 0.007 | -0.630 | 0.527 | 0.721 |
| Superior parietal | 8,655 | -0.009 | 0.008 | -1.110 | 0.267 | 0.558 |
| Supramarginal | 8,659 | -0.028 | 0.007 | -3.910 | <0.001 | <0.001 |
| Frontal pole | 8,662 | 0.003 | 0.007 | 0.370 | 0.709 | 0.838 |
| Insula | 8,662 | -0.008 | 0.007 | -1.090 | 0.275 | 0.558 |
| Superior temporal | 8,658 | -0.021 | 0.007 | -3.110 | 0.002 | 0.026 |
| Inferior frontal | 8,660 | -0.021 | 0.008 | -2.810 | 0.005 | 0.043 |
| Dorsal lateral prefrontal | 8,652 | -0.007 | 0.006 | -1.100 | 0.271 | 0.558 |
| Medial occipital | 8,658 | -0.009 | 0.009 | -1.010 | 0.313 | 0.581 |
| ***Cortical volume*** |  |  |  |  |  |  |
| Caudal anterior cingulate | 8,665 | -0.011 | 0.007 | -1.520 | 0.128 | 0.416 |
| Caudal middle frontal | 8,660 | 0.006 | 0.008 | 0.720 | 0.469 | 0.709 |
| Entorhinal | 8,662 | 0.011 | 0.008 | 1.310 | 0.191 | 0.440 |
| Fusiform | 8,661 | 0.001 | 0.007 | 0.080 | 0.934 | 0.971 |
| Inferior parietal | 8,663 | 0.010 | 0.008 | 1.290 | 0.197 | 0.440 |
| Inferior temporal | 8,662 | -0.009 | 0.007 | -1.230 | 0.220 | 0.440 |
| Isthmus cingulate | 8,661 | -0.005 | 0.008 | -0.670 | 0.504 | 0.709 |
| Lateral occipital | 8,660 | 0.010 | 0.008 | 1.270 | 0.203 | 0.440 |
| Lateral orbitofrontal | 8,659 | -0.012 | 0.007 | -1.570 | 0.117 | 0.416 |
| Medial orbitofrontal | 8,664 | -0.016 | 0.007 | -2.210 | 0.027 | 0.176 |
| Middle temporal | 8,658 | -0.006 | 0.007 | -0.890 | 0.373 | 0.647 |
| Parahippocampal | 8,663 | -0.010 | 0.009 | -1.110 | 0.268 | 0.498 |
| Paracentral | 8,662 | -0.004 | 0.008 | -0.500 | 0.614 | 0.798 |
| Postcentral | 8,663 | 0.006 | 0.008 | 0.820 | 0.412 | 0.670 |
| Posterior cingulate | 8,663 | -0.015 | 0.007 | -2.040 | 0.041 | 0.213 |
| Precentral | 8,659 | 0.005 | 0.008 | 0.650 | 0.518 | 0.709 |
| Precuneus | 8,655 | -0.003 | 0.007 | -0.400 | 0.688 | 0.806 |
| Rostral anterior cingulate | 8,664 | -0.010 | 0.007 | -1.410 | 0.160 | 0.440 |
| Superior parietal | 8,663 | -0.001 | 0.008 | -0.130 | 0.896 | 0.971 |
| Supramarginal | 8,664 | -0.021 | 0.007 | -2.930 | 0.003 | 0.039 |
| Frontal pole | 8,661 | 0.003 | 0.008 | 0.370 | 0.713 | 0.806 |
| Insula | 8,650 | -0.013 | 0.007 | -1.810 | 0.071 | 0.308 |
| Superior temporal | 8,650 | -0.018 | 0.007 | -2.530 | 0.011 | 0.095 |
| Inferior frontal | 8,656 | -0.026 | 0.008 | -3.390 | 0.001 | 0.026 |
| Dorsal lateral prefrontal | 8,657 | -0.003 | 0.006 | -0.460 | 0.646 | 0.800 |
| Medial occipital | 8,657 | <0.001 | 0.009 | <0.001 | 0.999 | 0.999 |
| ***Subcortical volumes*** |  |  |  |  |  |  |
| Thalamus | 8,644 | -0.008 | 0.007 | -1.190 | 0.233 | 0.513 |
| Caudate | 8,647 | 0.001 | 0.009 | 0.160 | 0.872 | 0.959 |
| Putamen | 8,635 | <0.001 | 0.008 | 0.030 | 0.975 | 0.975 |
| Pallidum | 8,654 | -0.005 | 0.008 | -0.660 | 0.507 | 0.697 |
| Hippocampus | 8,658 | -0.010 | 0.008 | -1.280 | 0.202 | 0.513 |
| Amygdala | 8,651 | -0.021 | 0.007 | -2.820 | 0.005 | 0.055 |
| Accumbens | 8,658 | -0.007 | 0.008 | -0.840 | 0.399 | 0.697 |
| left hippocampus | 8,639 | -0.017 | 0.008 | -2.040 | 0.042 | 0.154 |
| right hippocampus | 8,638 | -0.003 | 0.008 | -0.330 | 0.740 | 0.904 |
| left accumbens | 8,632 | 0.007 | 0.009 | 0.760 | 0.448 | 0.697 |
| right accumbens | 8,631 | -0.019 | 0.009 | -2.130 | 0.033 | 0.154 |

#### Table S5. The association between polygenic risk score for schizophrenia and brain structures. The model was conducted with age, age^2^, sex, total ICV, hemisphere, head position coordinates, genotype array, the first fifteen genetic principal components and birth weight, Townsend deprivation index, cannabis use and childhood traumatic events included as covariates.

| **Brain structure** | **N** | **β** | **SE** | **t** | **p** | **p_corrected_** |
| --- | --- | --- | --- | --- | --- | --- |
| ***Global*** |  |  |  |  |  |  |
| Cortical thickness | 8,622 | 0.002 | 0.009 | 0.210 | 0.831 | 0.831 |
| Surface area | 8,633 | -0.010 | 0.005 | -1.870 | 0.062 | 0.186 |
| Cortical volume | 8,637 | -0.004 | 0.006 | -0.750 | 0.456 | 0.684 |
| ***Lobes*** |  |  |  |  |  |  |
| ***Cortical thickness*** |  |  |  |  |  |  |
| Frontal lobe | 8,623 | -0.004 | 0.009 | -0.470 | 0.636 | 0.848 |
| Temporal lobe | 8,636 | 0.003 | 0.009 | 0.290 | 0.775 | 0.886 |
| Occipital lobe | 8,640 | 0.013 | 0.010 | 1.320 | 0.187 | 0.696 |
| Parietal lobe | 8,610 | 0.011 | 0.010 | 1.190 | 0.235 | 0.696 |
| Cingulate lobe | 8,642 | -0.009 | 0.009 | -1.070 | 0.285 | 0.696 |
| Insula lobe | 8,645 | -0.008 | 0.009 | -0.940 | 0.348 | 0.696 |
| Postcentral lobe | 8,643 | 0.006 | 0.009 | 0.680 | 0.500 | 0.800 |
| Paracentral lobe | 8,639 | 0.001 | 0.009 | 0.140 | 0.886 | 0.886 |
| ***Surface area*** |  |  |  |  |  |  |
| Frontal lobe | 8,634 | -0.007 | 0.006 | -1.180 | 0.238 | 0.382 |
| Temporal lobe | 8,634 | -0.016 | 0.006 | -2.690 | 0.007 | 0.056 |
| Occipital lobe | 8,632 | -0.005 | 0.008 | -0.550 | 0.582 | 0.619 |
| Parietal lobe | 8,636 | -0.010 | 0.007 | -1.470 | 0.143 | 0.382 |
| Cingulate lobe | 8,639 | -0.008 | 0.006 | -1.240 | 0.214 | 0.382 |
| Insula lobe | 8,641 | -0.008 | 0.007 | -1.180 | 0.239 | 0.382 |
| Postcentral lobe | 8,639 | -0.003 | 0.007 | -0.500 | 0.619 | 0.619 |
| Paracentral lobe | 8,639 | -0.005 | 0.007 | -0.710 | 0.479 | 0.619 |
| ***Cortical volume*** |  |  |  |  |  |  |
| Frontal lobe | 8,638 | -0.005 | 0.006 | -0.720 | 0.470 | 0.627 |
| Temporal lobe | 8,636 | -0.012 | 0.006 | -1.800 | 0.072 | 0.192 |
| Occipital lobe | 8,638 | 0.007 | 0.008 | 0.800 | 0.421 | 0.627 |
| Parietal lobe | 8,641 | -0.003 | 0.007 | -0.480 | 0.634 | 0.642 |
| Cingulate lobe | 8,639 | -0.014 | 0.007 | -1.960 | 0.051 | 0.192 |
| Insula lobe | 8,629 | -0.014 | 0.007 | -1.840 | 0.066 | 0.192 |
| Postcentral lobe | 8,642 | 0.006 | 0.008 | 0.810 | 0.419 | 0.627 |
| Paracentral lobe | 8,641 | -0.004 | 0.008 | -0.460 | 0.642 | 0.642 |
| ***Parcellations*** |  |  |  |  |  |  |
| ***Cortical thickness*** |  |  |  |  |  |  |
| Caudal anterior cingulate | 8,644 | -0.009 | 0.008 | -1.070 | 0.285 | 0.696 |
| Caudal middle frontal | 8,623 | 0.015 | 0.009 | 1.690 | 0.091 | 0.551 |
| Entorhinal | 8,632 | -0.005 | 0.009 | -0.570 | 0.570 | 0.780 |
| Fusiform | 8,638 | 0.011 | 0.009 | 1.240 | 0.216 | 0.696 |
| Inferior parietal | 8,622 | 0.009 | 0.009 | 0.940 | 0.346 | 0.696 |
| Inferior temporal | 8,642 | -0.002 | 0.009 | -0.180 | 0.859 | 0.938 |
| Isthmus cingulate | 8,642 | -0.002 | 0.009 | -0.170 | 0.862 | 0.938 |
| Lateral occipital | 8,637 | 0.012 | 0.010 | 1.210 | 0.226 | 0.696 |
| Lateral orbitofrontal | 8,638 | -0.021 | 0.009 | -2.260 | 0.024 | 0.551 |
| Medial orbitofrontal | 8,639 | -0.015 | 0.009 | -1.620 | 0.106 | 0.551 |
| Middle temporal | 8,643 | 0.001 | 0.009 | 0.120 | 0.902 | 0.938 |
| Parahippocampal | 8,643 | <0.001 | 0.009 | 0.050 | 0.964 | 0.964 |
| Paracentral | 8,639 | 0.001 | 0.009 | 0.140 | 0.886 | 0.938 |
| Postcentral | 8,643 | 0.006 | 0.009 | 0.680 | 0.500 | 0.722 |
| Posterior cingulate | 8,645 | -0.017 | 0.009 | -1.890 | 0.058 | 0.551 |
| Precentral | 8,631 | 0.007 | 0.009 | 0.730 | 0.466 | 0.722 |
| Precuneus | 8,629 | 0.007 | 0.010 | 0.770 | 0.442 | 0.722 |
| Rostral anterior cingulate | 8,642 | -0.002 | 0.008 | -0.270 | 0.790 | 0.938 |
| Superior parietal | 8,623 | 0.011 | 0.010 | 1.170 | 0.243 | 0.696 |
| Supramarginal | 8,624 | 0.011 | 0.009 | 1.190 | 0.235 | 0.696 |
| Frontal pole | 8,643 | -0.004 | 0.009 | -0.420 | 0.671 | 0.872 |
| Insula | 8,645 | -0.008 | 0.009 | -0.940 | 0.348 | 0.696 |
| Superior temporal | 8,634 | 0.008 | 0.010 | 0.830 | 0.404 | 0.722 |
| Inferior frontal | 8,622 | -0.016 | 0.009 | -1.740 | 0.082 | 0.551 |
| Dorsal lateral prefrontal | 8,613 | 0.006 | 0.009 | 0.680 | 0.498 | 0.722 |
| Medial occipital | 8,639 | 0.010 | 0.010 | 1.040 | 0.297 | 0.696 |
| ***Surface area*** |  |  |  |  |  |  |
| Caudal anterior cingulate | 8,640 | -0.008 | 0.007 | -1.130 | 0.260 | 0.576 |
| Caudal middle frontal | 8,637 | <0.001 | 0.008 | -0.050 | 0.961 | 0.975 |
| Entorhinal | 8,646 | 0.007 | 0.009 | 0.780 | 0.436 | 0.655 |
| Fusiform | 8,638 | -0.008 | 0.007 | -1.210 | 0.228 | 0.576 |
| Inferior parietal | 8,641 | 0.006 | 0.007 | 0.850 | 0.393 | 0.639 |
| Inferior temporal | 8,638 | -0.013 | 0.007 | -1.800 | 0.072 | 0.374 |
| Isthmus cingulate | 8,636 | -0.006 | 0.007 | -0.880 | 0.380 | 0.639 |
| Lateral occipital | 8,636 | 0.002 | 0.008 | 0.230 | 0.822 | 0.929 |
| Lateral orbitofrontal | 8,640 | -0.001 | 0.007 | -0.100 | 0.923 | 0.975 |
| Medial orbitofrontal | 8,640 | -0.004 | 0.006 | -0.570 | 0.566 | 0.736 |
| Middle temporal | 8,632 | -0.013 | 0.007 | -1.800 | 0.072 | 0.374 |
| Parahippocampal | 8,641 | -0.010 | 0.008 | -1.330 | 0.183 | 0.576 |
| Paracentral | 8,639 | -0.005 | 0.007 | -0.710 | 0.479 | 0.655 |
| Postcentral | 8,639 | -0.003 | 0.007 | -0.500 | 0.619 | 0.766 |
| Posterior cingulate | 8,639 | -0.010 | 0.007 | -1.470 | 0.143 | 0.576 |
| Precentral | 8,637 | <0.001 | 0.007 | 0.030 | 0.975 | 0.975 |
| Precuneus | 8,634 | -0.008 | 0.007 | -1.060 | 0.288 | 0.576 |
| Rostral anterior cingulate | 8,639 | -0.005 | 0.007 | -0.720 | 0.474 | 0.655 |
| Superior parietal | 8,634 | -0.008 | 0.008 | -1.070 | 0.286 | 0.576 |
| Supramarginal | 8,638 | -0.028 | 0.007 | -3.910 | <0.001 | <0.001 |
| Frontal pole | 8,641 | 0.003 | 0.007 | 0.410 | 0.682 | 0.806 |
| Insula | 8,641 | -0.008 | 0.007 | -1.180 | 0.239 | 0.576 |
| Superior temporal | 8,637 | -0.021 | 0.007 | -3.090 | 0.002 | 0.026 |
| Inferior frontal | 8,639 | -0.022 | 0.008 | -2.880 | 0.004 | 0.035 |
| Dorsal lateral prefrontal | 8,631 | -0.007 | 0.006 | -1.080 | 0.281 | 0.576 |
| Medial occipital | 8,637 | -0.009 | 0.009 | -0.950 | 0.344 | 0.639 |
| ***Cortical volume*** |  |  |  |  |  |  |
| Caudal anterior cingulate | 8,644 | -0.011 | 0.007 | -1.500 | 0.135 | 0.422 |
| Caudal middle frontal | 8,639 | 0.005 | 0.008 | 0.650 | 0.517 | 0.707 |
| Entorhinal | 8,641 | 0.010 | 0.008 | 1.240 | 0.214 | 0.428 |
| Fusiform | 8,640 | 0.001 | 0.007 | 0.090 | 0.928 | 0.977 |
| Inferior parietal | 8,642 | 0.010 | 0.008 | 1.330 | 0.183 | 0.428 |
| Inferior temporal | 8,641 | -0.009 | 0.007 | -1.250 | 0.211 | 0.428 |
| Isthmus cingulate | 8,640 | -0.006 | 0.008 | -0.740 | 0.459 | 0.663 |
| Lateral occipital | 8,639 | 0.010 | 0.008 | 1.280 | 0.202 | 0.428 |
| Lateral orbitofrontal | 8,638 | -0.011 | 0.007 | -1.450 | 0.146 | 0.422 |
| Medial orbitofrontal | 8,644 | -0.014 | 0.007 | -1.990 | 0.046 | 0.239 |
| Middle temporal | 8,637 | -0.005 | 0.007 | -0.750 | 0.452 | 0.663 |
| Parahippocampal | 8,642 | -0.009 | 0.009 | -1.080 | 0.282 | 0.524 |
| Paracentral | 8,641 | -0.004 | 0.008 | -0.460 | 0.642 | 0.796 |
| Postcentral | 8,642 | 0.006 | 0.008 | 0.810 | 0.419 | 0.663 |
| Posterior cingulate | 8,642 | -0.015 | 0.007 | -2.000 | 0.045 | 0.239 |
| Precentral | 8,638 | 0.006 | 0.008 | 0.750 | 0.453 | 0.663 |
| Precuneus | 8,634 | -0.002 | 0.007 | -0.330 | 0.742 | 0.839 |
| Rostral anterior cingulate | 8,643 | -0.011 | 0.007 | -1.460 | 0.144 | 0.422 |
| Superior parietal | 8,642 | -0.001 | 0.008 | -0.080 | 0.939 | 0.977 |
| Supramarginal | 8,643 | -0.020 | 0.007 | -2.820 | 0.005 | 0.065 |
| Frontal pole | 8,640 | 0.004 | 0.008 | 0.460 | 0.643 | 0.796 |
| Insula | 8,629 | -0.014 | 0.007 | -1.840 | 0.066 | 0.286 |
| Superior temporal | 8,629 | -0.018 | 0.007 | -2.420 | 0.015 | 0.130 |
| Inferior frontal | 8,635 | -0.026 | 0.008 | -3.330 | 0.001 | 0.026 |
| Dorsal lateral prefrontal | 8,637 | -0.002 | 0.006 | -0.350 | 0.729 | 0.839 |
| Medial occipital | 8,636 | <0.001 | 0.009 | 0.020 | 0.984 | 0.984 |
| ***Subcortical volumes*** |  |  |  |  |  |  |
| Thalamus | 8,623 | -0.008 | 0.007 | -1.210 | 0.226 | 0.497 |
| Caudate | 8,626 | 0.002 | 0.009 | 0.190 | 0.849 | 0.890 |
| Putamen | 8,614 | -0.001 | 0.008 | -0.140 | 0.890 | 0.890 |
| Pallidum | 8,633 | -0.005 | 0.008 | -0.670 | 0.502 | 0.690 |
| Hippocampus | 8,637 | -0.010 | 0.008 | -1.280 | 0.200 | 0.497 |
| Amygdala | 8,630 | -0.021 | 0.007 | -2.820 | 0.005 | 0.055 |
| Accumbens | 8,637 | -0.006 | 0.008 | -0.690 | 0.493 | 0.690 |
| Left hippocampus | 8,618 | -0.017 | 0.008 | -2.040 | 0.041 | 0.194 |
| Right hippocampus | 8,617 | -0.003 | 0.009 | -0.330 | 0.739 | 0.890 |
| Left accumbens | 8,611 | 0.007 | 0.009 | 0.840 | 0.400 | 0.690 |
| Right accumbens | 8,610 | -0.017 | 0.009 | -1.940 | 0.053 | 0.194 |

#### Table S6. The interaction between polygenic risk score for schizophrenia and childhood traumatic events on brain structures.

| **Brain structure** | **N** | **β** | **SE** | **t** | **p_uncorrected_** | **p_corrected_** |
| --- | --- | --- | --- | --- | --- | --- |
| ***Global*** |  |  |  |  |  |  |
| Cortical thickness | 12,963 | 0.001 | 0.003 | 0.400 | 0.687 | 0.801 |
| Surface area | 12,985 | <0.001 | 0.002 | 0.250 | 0.801 | 0.801 |
| Cortical volume | 12,990 | 0.002 | 0.002 | 1.120 | 0.262 | 0.786 |
| ***Lobes*** |  |  |  |  |  |  |
| ***Cortical thickness*** |  |  |  |  |  |  |
| Frontal lobe | 12,965 | 0.002 | 0.003 | 0.750 | 0.454 | 0.979 |
| Temporal lobe | 12,980 | -0.003 | 0.003 | -0.820 | 0.414 | 0.979 |
| Occipital lobe | 12,994 | 0.001 | 0.003 | 0.330 | 0.745 | 0.979 |
| Parietal lobe | 12,947 | <0.001 | 0.003 | 0.030 | 0.979 | 0.979 |
| Cingulate lobe | 12,997 | 0.002 | 0.003 | 0.510 | 0.613 | 0.979 |
| Insula lobe | 12,999 | <0.001 | 0.003 | -0.130 | 0.899 | 0.979 |
| Postcentral lobe | 12,992 | 0.004 | 0.003 | 1.170 | 0.242 | 0.979 |
| Paracentral lobe | 12,990 | 0.002 | 0.003 | 0.530 | 0.596 | 0.979 |
| ***Surface area*** |  |  |  |  |  |  |
| Frontal lobe | 12,984 | -0.001 | 0.002 | -0.610 | 0.544 | 0.635 |
| Temporal lobe | 12,985 | 0.002 | 0.002 | 0.900 | 0.368 | 0.635 |
| Occipital lobe | 12,984 | 0.001 | 0.003 | 0.330 | 0.742 | 0.742 |
| Parietal lobe | 12,989 | 0.002 | 0.002 | 0.720 | 0.473 | 0.635 |
| Cingulate lobe | 12,991 | -0.001 | 0.002 | -0.590 | 0.556 | 0.635 |
| Insula lobe | 12,993 | 0.002 | 0.002 | 0.820 | 0.413 | 0.635 |
| Postcentral lobe | 12,990 | 0.002 | 0.002 | 0.840 | 0.398 | 0.635 |
| Paracentral lobe | 12,990 | 0.004 | 0.003 | 1.520 | 0.128 | 0.635 |
| ***Cortical volume*** |  |  |  |  |  |  |
| Frontal lobe | 12,989 | 0.001 | 0.002 | 0.470 | 0.635 | 0.726 |
| Temporal lobe | 12,989 | 0.002 | 0.002 | 0.740 | 0.459 | 0.659 |
| Occipital lobe | 12,991 | 0.002 | 0.003 | 0.680 | 0.494 | 0.659 |
| Parietal lobe | 12,992 | 0.003 | 0.002 | 1.230 | 0.219 | 0.584 |
| Cingulate lobe | 12,993 | <0.001 | 0.002 | 0.130 | 0.893 | 0.893 |
| Insula lobe | 12,979 | 0.003 | 0.003 | 0.960 | 0.336 | 0.659 |
| Postcentral lobe | 12,994 | 0.005 | 0.003 | 1.780 | 0.075 | 0.372 |
| Paracentral lobe | 12,996 | 0.005 | 0.003 | 1.680 | 0.093 | 0.372 |
| ***Parcellations*** |  |  |  |  |  |  |
| ***Cortical thickness*** |  |  |  |  |  |  |
| Caudal anterior cingulate | 12,999 | 0.002 | 0.003 | 0.650 | 0.517 | 0.962 |
| Caudal middle frontal | 12,966 | 0.003 | 0.003 | 0.840 | 0.400 | 0.962 |
| Entorhinal | 12,974 | -0.004 | 0.003 | -1.230 | 0.219 | 0.962 |
| Fusiform | 12,986 | -0.001 | 0.003 | -0.210 | 0.837 | 0.962 |
| Inferior parietal | 12,961 | -0.002 | 0.003 | -0.480 | 0.628 | 0.962 |
| Inferior temporal | 12,995 | -0.001 | 0.003 | -0.360 | 0.720 | 0.962 |
| Isthmus cingulate | 12,997 | 0.001 | 0.003 | 0.440 | 0.658 | 0.962 |
| Lateral occipital | 12,992 | 0.003 | 0.003 | 0.860 | 0.391 | 0.962 |
| Lateral orbitofrontal | 12,988 | 0.001 | 0.003 | 0.380 | 0.706 | 0.962 |
| Medial orbitofrontal | 12,989 | 0.005 | 0.003 | 1.560 | 0.119 | 0.962 |
| Middle temporal | 12,998 | -0.002 | 0.003 | -0.630 | 0.527 | 0.962 |
| Parahippocampal | 12,995 | -0.003 | 0.003 | -0.920 | 0.360 | 0.962 |
| Paracentral | 12,990 | 0.002 | 0.003 | 0.530 | 0.596 | 0.962 |
| Postcentral | 12,992 | 0.004 | 0.003 | 1.170 | 0.242 | 0.962 |
| Posterior cingulate | 12,999 | 0.001 | 0.003 | 0.290 | 0.772 | 0.962 |
| Precentral | 12,975 | 0.002 | 0.003 | 0.660 | 0.511 | 0.962 |
| Precuneus | 12,977 | 0.001 | 0.003 | 0.170 | 0.867 | 0.962 |
| Rostral anterior cingulate | 12,995 | -0.001 | 0.003 | -0.210 | 0.832 | 0.962 |
| Superior parietal | 12,966 | 0.001 | 0.003 | 0.220 | 0.824 | 0.962 |
| Supramarginal | 12,964 | 0.001 | 0.003 | 0.220 | 0.827 | 0.962 |
| Frontal pole | 12,993 | 0.001 | 0.003 | 0.290 | 0.769 | 0.962 |
| Insula | 12,999 | <0.001 | 0.003 | -0.130 | 0.899 | 0.962 |
| Superior temporal | 12,978 | <0.001 | 0.003 | 0.050 | 0.961 | 0.962 |
| Inferior frontal | 12,964 | <0.001 | 0.003 | 0.050 | 0.962 | 0.962 |
| Dorsal lateral prefrontal | 12,940 | 0.001 | 0.003 | 0.360 | 0.717 | 0.962 |
| Medial occipital | 12,990 | <0.001 | 0.003 | -0.090 | 0.930 | 0.962 |
| ***Surface area*** |  |  |  |  |  |  |
| Caudal anterior cingulate | 12,993 | <0.001 | 0.002 | -0.150 | 0.879 | 0.941 |
| Caudal middle frontal | 12,989 | 0.002 | 0.003 | 0.870 | 0.383 | 0.785 |
| Entorhinal | 13,000 | 0.003 | 0.003 | 0.980 | 0.329 | 0.785 |
| Fusiform | 12,991 | 0.001 | 0.002 | 0.460 | 0.645 | 0.883 |
| Inferior parietal | 12,996 | 0.003 | 0.003 | 1.150 | 0.250 | 0.785 |
| Inferior temporal | 12,992 | 0.001 | 0.002 | 0.400 | 0.690 | 0.897 |
| Isthmus cingulate | 12,988 | -0.002 | 0.003 | -0.650 | 0.513 | 0.785 |
| Lateral occipital | 12,988 | 0.002 | 0.003 | 0.880 | 0.381 | 0.785 |
| Lateral orbitofrontal | 12,993 | -0.002 | 0.003 | -0.880 | 0.376 | 0.785 |
| Medial orbitofrontal | 12,992 | -0.006 | 0.002 | -2.500 | 0.013 | 0.338 |
| Middle temporal | 12,985 | <0.001 | 0.002 | 0.100 | 0.921 | 0.941 |
| Parahippocampal | 12,994 | 0.003 | 0.003 | 1.290 | 0.197 | 0.785 |
| Paracentral | 12,990 | 0.004 | 0.003 | 1.520 | 0.128 | 0.785 |
| Postcentral | 12,990 | 0.002 | 0.002 | 0.840 | 0.398 | 0.785 |
| Posterior cingulate | 12,988 | -0.001 | 0.002 | -0.350 | 0.727 | 0.900 |
| Precentral | 12,992 | 0.003 | 0.002 | 1.040 | 0.300 | 0.785 |
| Precuneus | 12,982 | <0.001 | 0.003 | 0.170 | 0.864 | 0.941 |
| Rostral anterior cingulate | 12,991 | <0.001 | 0.002 | -0.070 | 0.941 | 0.941 |
| Superior parietal | 12,986 | 0.004 | 0.003 | 1.300 | 0.193 | 0.785 |
| Supramarginal | 12,991 | -0.002 | 0.002 | -0.720 | 0.472 | 0.785 |
| Frontal pole | 12,997 | -0.002 | 0.003 | -0.680 | 0.500 | 0.785 |
| Insula | 12,993 | 0.002 | 0.002 | 0.820 | 0.413 | 0.785 |
| Superior temporal | 12,991 | 0.001 | 0.002 | 0.520 | 0.605 | 0.874 |
| Inferior frontal | 12,989 | 0.002 | 0.003 | 0.740 | 0.458 | 0.785 |
| Dorsal lateral prefrontal | 12,983 | -0.003 | 0.002 | -1.300 | 0.193 | 0.785 |
| Medial occipital | 12,986 | -0.001 | 0.003 | -0.300 | 0.762 | 0.901 |
| ***Cortical volume*** |  |  |  |  |  |  |
| Caudal anterior cingulate | 13,000 | <0.001 | 0.003 | 0.110 | 0.912 | 0.930 |
| Caudal middle frontal | 12,992 | 0.004 | 0.003 | 1.570 | 0.116 | 0.603 |
| Entorhinal | 12,995 | 0.002 | 0.003 | 0.680 | 0.494 | 0.872 |
| Fusiform | 12,995 | 0.001 | 0.003 | 0.540 | 0.592 | 0.872 |
| Inferior parietal | 12,995 | 0.003 | 0.003 | 1.080 | 0.279 | 0.872 |
| Inferior temporal | 12,995 | 0.002 | 0.003 | 0.610 | 0.544 | 0.872 |
| Isthmus cingulate | 12,994 | <0.001 | 0.003 | -0.170 | 0.868 | 0.930 |
| Lateral occipital | 12,992 | 0.004 | 0.003 | 1.420 | 0.157 | 0.680 |
| Lateral orbitofrontal | 12,988 | -0.001 | 0.003 | -0.440 | 0.658 | 0.872 |
| Medial orbitofrontal | 12,996 | -0.002 | 0.003 | -0.710 | 0.478 | 0.872 |
| Middle temporal | 12,990 | -0.001 | 0.003 | -0.420 | 0.673 | 0.872 |
| Parahippocampal | 12,997 | 0.002 | 0.003 | 0.630 | 0.527 | 0.872 |
| Paracentral | 12,996 | 0.005 | 0.003 | 1.680 | 0.093 | 0.603 |
| Postcentral | 12,994 | 0.005 | 0.003 | 1.780 | 0.075 | 0.603 |
| Posterior cingulate | 12,994 | <0.001 | 0.003 | 0.120 | 0.908 | 0.930 |
| Precentral | 12,989 | 0.005 | 0.003 | 1.780 | 0.075 | 0.603 |
| Precuneus | 12,979 | 0.002 | 0.003 | 0.860 | 0.392 | 0.872 |
| Rostral anterior cingulate | 12,998 | 0.001 | 0.003 | 0.380 | 0.704 | 0.872 |
| Superior parietal | 12,995 | 0.005 | 0.003 | 1.890 | 0.059 | 0.603 |
| Supramarginal | 12,995 | -0.001 | 0.003 | -0.520 | 0.603 | 0.872 |
| Frontal pole | 12,988 | 0.001 | 0.003 | 0.270 | 0.787 | 0.930 |
| Insula | 12,979 | 0.003 | 0.003 | 0.960 | 0.336 | 0.872 |
| Superior temporal | 12,980 | 0.002 | 0.003 | 0.780 | 0.437 | 0.872 |
| Inferior frontal | 12,987 | 0.003 | 0.003 | 0.940 | 0.345 | 0.872 |
| Dorsal lateral prefrontal | 12,989 | -0.001 | 0.002 | -0.430 | 0.668 | 0.872 |
| Medial occipital | 12,988 | <0.001 | 0.003 | -0.090 | 0.930 | 0.930 |
| ***Subcortical volumes*** |  |  |  |  |  |  |
| Thalamus | 12,972 | 0.001 | 0.002 | 0.520 | 0.605 | 0.847 |
| Caudate | 12,960 | 0.001 | 0.003 | 0.360 | 0.716 | 0.847 |
| Putamen | 12,958 | -0.001 | 0.003 | -0.510 | 0.607 | 0.847 |
| Pallidum | 12,981 | 0.003 | 0.003 | 1.100 | 0.270 | 0.847 |
| Hippocampus | 12,984 | 0.001 | 0.003 | 0.460 | 0.645 | 0.847 |
| Amygdala | 12,983 | 0.002 | 0.003 | 0.690 | 0.491 | 0.847 |
| Accumbens | 12,985 | -0.002 | 0.003 | -0.560 | 0.576 | 0.847 |
| Left hippocampus | 12,952 | 0.002 | 0.003 | 0.760 | 0.447 | 0.847 |
| Right hippocampus | 12,949 | <0.001 | 0.003 | 0.010 | 0.995 | 0.995 |
| Left accumbens | 12,940 | -0.001 | 0.003 | -0.290 | 0.770 | 0.847 |
| Right accumbens | 12,942 | -0.002 | 0.003 | -0.620 | 0.534 | 0.847 |

#### Table S7. The interaction between polygenic risk score for schizophrenia and cannabis use (never/ever) on brain structures.

| **Brain structure** | **N** | **β** | **SE** | **z** | **p_uncorrected_** | **p_corrected_** |
| --- | --- | --- | --- | --- | --- | --- |
| ***Global*** |  |  |  |  |  |  |
| Cortical thickness | 13,215 | 0.025 | 0.019 | 1.290 | 0.196 | 0.294 |
| Surface area | 13,237 | 0.003 | 0.011 | 0.300 | 0.763 | 0.763 |
| Cortical volume | 13,242 | 0.019 | 0.012 | 1.650 | 0.098 | 0.294 |
| ***Lobes*** |  |  |  |  |  |  |
| ***Cortical thickness*** |  |  |  |  |  |  |
| Frontal lobe | 13,217 | 0.041 | 0.019 | 2.140 | 0.032 | 0.148 |
| Temporal lobe | 13,230 | 0.017 | 0.019 | 0.910 | 0.362 | 0.579 |
| Occipital lobe | 13,246 | -0.001 | 0.019 | -0.080 | 0.939 | 0.939 |
| Parietal lobe | 13,198 | 0.014 | 0.019 | 0.730 | 0.468 | 0.624 |
| Cingulate lobe | 13,249 | -0.002 | 0.017 | -0.130 | 0.898 | 0.939 |
| Insula lobe | 13,251 | 0.038 | 0.018 | 2.090 | 0.037 | 0.148 |
| Postcentral lobe | 13,244 | 0.022 | 0.019 | 1.150 | 0.251 | 0.502 |
| Paracentral lobe | 13,242 | 0.023 | 0.019 | 1.220 | 0.224 | 0.502 |
| ***Surface area*** |  |  |  |  |  |  |
| Frontal lobe | 13,236 | 0.004 | 0.012 | 0.300 | 0.765 | 0.765 |
| Temporal lobe | 13,236 | 0.006 | 0.012 | 0.470 | 0.638 | 0.729 |
| Occipital lobe | 13,235 | -0.016 | 0.017 | -0.940 | 0.348 | 0.696 |
| Parietal lobe | 13,241 | 0.016 | 0.013 | 1.210 | 0.228 | 0.696 |
| Cingulate lobe | 13,243 | 0.010 | 0.013 | 0.740 | 0.461 | 0.729 |
| Insula lobe | 13,245 | -0.007 | 0.014 | -0.510 | 0.610 | 0.729 |
| Postcentral lobe | 13,242 | 0.015 | 0.014 | 1.070 | 0.287 | 0.696 |
| Paracentral lobe | 13,242 | 0.021 | 0.015 | 1.410 | 0.158 | 0.696 |
| ***Cortical volume*** |  |  |  |  |  |  |
| Frontal lobe | 13,241 | 0.022 | 0.013 | 1.720 | 0.085 | 0.170 |
| Temporal lobe | 13,241 | 0.018 | 0.013 | 1.410 | 0.158 | 0.253 |
| Occipital lobe | 13,243 | -0.017 | 0.016 | -1.050 | 0.294 | 0.319 |
| Parietal lobe | 13,244 | 0.025 | 0.014 | 1.820 | 0.069 | 0.170 |
| Cingulate lobe | 13,245 | 0.017 | 0.014 | 1.240 | 0.216 | 0.288 |
| Insula lobe | 13,231 | 0.015 | 0.015 | 1.000 | 0.319 | 0.319 |
| Postcentral lobe | 13,246 | 0.026 | 0.015 | 1.740 | 0.081 | 0.170 |
| Paracentral lobe | 13,248 | 0.040 | 0.016 | 2.520 | 0.012 | 0.096 |
| ***Parcellations*** |  |  |  |  |  |  |
| ***Cortical thickness*** |  |  |  |  |  |  |
| Caudal anterior cingulate | 13,251 | -0.010 | 0.016 | -0.600 | 0.546 | 0.676 |
| Caudal middle frontal | 13,218 | 0.038 | 0.018 | 2.060 | 0.039 | 0.308 |
| Entorhinal | 13,225 | -0.002 | 0.018 | -0.130 | 0.899 | 0.935 |
| Fusiform | 13,238 | 0.013 | 0.018 | 0.730 | 0.468 | 0.640 |
| Inferior parietal | 13,213 | <0.001 | 0.019 | 0.020 | 0.988 | 0.988 |
| Inferior temporal | 13,247 | 0.025 | 0.018 | 1.360 | 0.173 | 0.346 |
| Isthmus cingulate | 13,249 | -0.013 | 0.018 | -0.730 | 0.464 | 0.640 |
| Lateral occipital | 13,244 | 0.017 | 0.019 | 0.900 | 0.368 | 0.563 |
| Lateral orbitofrontal | 13,240 | 0.007 | 0.019 | 0.360 | 0.718 | 0.812 |
| Medial orbitofrontal | 13,241 | 0.034 | 0.018 | 1.840 | 0.066 | 0.308 |
| Middle temporal | 13,250 | 0.027 | 0.018 | 1.480 | 0.138 | 0.346 |
| Parahippocampal | 13,246 | 0.013 | 0.019 | 0.680 | 0.498 | 0.647 |
| Paracentral | 13,242 | 0.023 | 0.019 | 1.220 | 0.224 | 0.416 |
| Postcentral | 13,244 | 0.022 | 0.019 | 1.150 | 0.251 | 0.435 |
| Posterior cingulate | 13,251 | 0.004 | 0.018 | 0.210 | 0.835 | 0.905 |
| Precentral | 13,226 | 0.034 | 0.019 | 1.800 | 0.071 | 0.308 |
| Precuneus | 13,229 | 0.031 | 0.019 | 1.620 | 0.106 | 0.346 |
| Rostral anterior cingulate | 13,247 | 0.031 | 0.017 | 1.840 | 0.066 | 0.308 |
| Superior parietal | 13,218 | 0.010 | 0.019 | 0.500 | 0.619 | 0.732 |
| Supramarginal | 13,216 | 0.017 | 0.018 | 0.950 | 0.343 | 0.557 |
| Frontal pole | 13,245 | 0.037 | 0.017 | 2.140 | 0.032 | 0.308 |
| Insula | 13,251 | 0.038 | 0.018 | 2.090 | 0.037 | 0.308 |
| Superior temporal | 13,230 | 0.028 | 0.019 | 1.460 | 0.145 | 0.346 |
| Inferior frontal | 13,216 | 0.025 | 0.019 | 1.360 | 0.173 | 0.346 |
| Dorsal lateral prefrontal | 13,190 | 0.026 | 0.019 | 1.400 | 0.162 | 0.346 |
| Medial occipital | 13,242 | -0.027 | 0.019 | -1.400 | 0.162 | 0.346 |
| ***Surface area*** |  |  |  |  |  |  |
| Caudal anterior cingulate | 13,245 | -0.011 | 0.014 | -0.800 | 0.425 | 0.801 |
| Caudal middle frontal | 13,241 | -0.001 | 0.015 | -0.060 | 0.951 | 0.951 |
| Entorhinal | 13,252 | -0.014 | 0.017 | -0.800 | 0.423 | 0.801 |
| Fusiform | 13,243 | 0.001 | 0.014 | 0.070 | 0.944 | 0.951 |
| Inferior parietal | 13,248 | 0.011 | 0.015 | 0.730 | 0.462 | 0.801 |
| Inferior temporal | 13,244 | 0.006 | 0.014 | 0.390 | 0.696 | 0.827 |
| Isthmus cingulate | 13,240 | 0.011 | 0.014 | 0.780 | 0.438 | 0.801 |
| Lateral occipital | 13,240 | -0.019 | 0.016 | -1.220 | 0.221 | 0.801 |
| Lateral orbitofrontal | 13,245 | 0.013 | 0.014 | 0.880 | 0.379 | 0.801 |
| Medial orbitofrontal | 13,244 | 0.013 | 0.013 | 1.010 | 0.315 | 0.801 |
| Middle temporal | 13,237 | 0.017 | 0.014 | 1.210 | 0.228 | 0.801 |
| Parahippocampal | 13,246 | 0.007 | 0.015 | 0.440 | 0.662 | 0.827 |
| Paracentral | 13,242 | 0.021 | 0.015 | 1.410 | 0.158 | 0.801 |
| Postcentral | 13,242 | 0.015 | 0.014 | 1.070 | 0.287 | 0.801 |
| Posterior cingulate | 13,240 | 0.029 | 0.014 | 2.080 | 0.037 | 0.801 |
| Precentral | 13,244 | 0.016 | 0.014 | 1.120 | 0.264 | 0.801 |
| Precuneus | 13,233 | 0.001 | 0.015 | 0.070 | 0.943 | 0.951 |
| Rostral anterior cingulate | 13,242 | -0.004 | 0.014 | -0.270 | 0.785 | 0.887 |
| Superior parietal | 13,238 | 0.017 | 0.016 | 1.090 | 0.274 | 0.801 |
| Supramarginal | 13,242 | 0.014 | 0.014 | 0.980 | 0.329 | 0.801 |
| Frontal pole | 13,249 | 0.009 | 0.015 | 0.610 | 0.540 | 0.827 |
| Insula | 13,245 | -0.007 | 0.014 | -0.510 | 0.610 | 0.827 |
| Superior temporal | 13,242 | 0.011 | 0.014 | 0.820 | 0.410 | 0.801 |
| Inferior frontal | 13,241 | -0.007 | 0.015 | -0.490 | 0.621 | 0.827 |
| Dorsal lateral prefrontal | 13,235 | -0.006 | 0.013 | -0.510 | 0.608 | 0.827 |
| Medial occipital | 13,238 | -0.007 | 0.018 | -0.380 | 0.700 | 0.827 |
| ***Cortical volume*** |  |  |  |  |  |  |
| Caudal anterior cingulate | 13,252 | -0.003 | 0.015 | -0.230 | 0.821 | 0.854 |
| Caudal middle frontal | 13,244 | 0.021 | 0.015 | 1.340 | 0.179 | 0.388 |
| Entorhinal | 13,247 | -0.013 | 0.017 | -0.770 | 0.441 | 0.609 |
| Fusiform | 13,247 | 0.007 | 0.015 | 0.500 | 0.619 | 0.768 |
| Inferior parietal | 13,247 | 0.012 | 0.015 | 0.760 | 0.445 | 0.609 |
| Inferior temporal | 13,246 | 0.015 | 0.014 | 1.070 | 0.285 | 0.494 |
| Isthmus cingulate | 13,246 | 0.004 | 0.015 | 0.260 | 0.794 | 0.854 |
| Lateral occipital | 13,244 | -0.002 | 0.015 | -0.150 | 0.877 | 0.877 |
| Lateral orbitofrontal | 13,240 | 0.020 | 0.015 | 1.370 | 0.171 | 0.388 |
| Medial orbitofrontal | 13,248 | 0.028 | 0.015 | 1.900 | 0.058 | 0.301 |
| Middle temporal | 13,242 | 0.019 | 0.014 | 1.300 | 0.195 | 0.390 |
| Parahippocampal | 13,249 | 0.008 | 0.018 | 0.460 | 0.643 | 0.768 |
| Paracentral | 13,248 | 0.040 | 0.016 | 2.520 | 0.012 | 0.156 |
| Postcentral | 13,246 | 0.026 | 0.015 | 1.740 | 0.081 | 0.301 |
| Posterior cingulate | 13,246 | 0.032 | 0.015 | 2.130 | 0.034 | 0.247 |
| Precentral | 13,241 | 0.041 | 0.015 | 2.670 | 0.008 | 0.156 |
| Precuneus | 13,231 | 0.017 | 0.015 | 1.110 | 0.266 | 0.494 |
| Rostral anterior cingulate | 13,250 | 0.007 | 0.015 | 0.450 | 0.650 | 0.768 |
| Superior parietal | 13,247 | 0.023 | 0.016 | 1.450 | 0.146 | 0.388 |
| Supramarginal | 13,246 | 0.026 | 0.015 | 1.800 | 0.072 | 0.301 |
| Frontal pole | 13,239 | 0.034 | 0.016 | 2.080 | 0.038 | 0.247 |
| Insula | 13,231 | 0.015 | 0.015 | 1.000 | 0.319 | 0.518 |
| Superior temporal | 13,232 | 0.023 | 0.015 | 1.590 | 0.111 | 0.361 |
| Inferior frontal | 13,239 | 0.006 | 0.015 | 0.400 | 0.691 | 0.781 |
| Dorsal lateral prefrontal | 13,241 | 0.011 | 0.013 | 0.810 | 0.417 | 0.609 |
| Medial occipital | 13,239 | -0.024 | 0.018 | -1.350 | 0.178 | 0.388 |
| ***Subcortical volumes*** |  |  |  |  |  |  |
| Thalamus | 13,224 | 0.001 | 0.014 | 0.070 | 0.945 | 0.993 |
| Caudate | 13,212 | -0.004 | 0.017 | -0.240 | 0.812 | 0.993 |
| Putamen | 13,210 | 0.003 | 0.017 | 0.170 | 0.866 | 0.993 |
| Pallidum | 13,233 | <0.001 | 0.016 | -0.010 | 0.993 | 0.993 |
| Hippocampus | 13,235 | 0.012 | 0.016 | 0.760 | 0.449 | 0.792 |
| Amygdala | 13,235 | 0.031 | 0.015 | 2.040 | 0.041 | 0.451 |
| Accumbens | 13,237 | 0.021 | 0.016 | 1.300 | 0.194 | 0.711 |
| Left hippocampus | 13,203 | 0.011 | 0.017 | 0.670 | 0.504 | 0.792 |
| Right hippocampus | 13,200 | 0.013 | 0.017 | 0.730 | 0.463 | 0.792 |
| Left accumbens | 13,193 | 0.027 | 0.018 | 1.520 | 0.129 | 0.710 |
| Right accumbens | 13,193 | 0.017 | 0.018 | 0.950 | 0.341 | 0.792 |

#### Table S8. The interaction between polygenic risk score for schizophrenia and birth weight on brain structures.

| **Brain structure** | **N** | **β** | **SE** | **z** | **p_uncorrected_** | **p_corrected_** |
| --- | --- | --- | --- | --- | --- | --- |
| ***Global*** |  |  |  |  |  |  |
| Cortical thickness | 11,889 | 0.006 | 0.013 | 0.440 | 0.660 | 0.885 |
| Surface area | 11,896 | -0.004 | 0.007 | -0.610 | 0.543 | 0.885 |
| Cortical volume | 11,907 | 0.001 | 0.008 | 0.140 | 0.885 | 0.885 |
| ***Lobes*** |  |  |  |  |  |  |
| ***Cortical thickness*** |  |  |  |  |  |  |
| Frontal lobe | 11,891 | 0.010 | 0.013 | 0.750 | 0.452 | 0.853 |
| Temporal lobe | 11,908 | 0.011 | 0.013 | 0.900 | 0.366 | 0.853 |
| Occipital lobe | 11,914 | -0.006 | 0.013 | -0.430 | 0.664 | 0.853 |
| Parietal lobe | 11,871 | 0.002 | 0.013 | 0.190 | 0.853 | 0.853 |
| Cingulate lobe | 11,916 | 0.008 | 0.012 | 0.660 | 0.512 | 0.853 |
| Insula lobe | 11,920 | 0.005 | 0.012 | 0.430 | 0.669 | 0.853 |
| Postcentral lobe | 11,918 | -0.003 | 0.013 | -0.260 | 0.797 | 0.853 |
| Paracentral lobe | 11,912 | 0.018 | 0.013 | 1.410 | 0.158 | 0.853 |
| ***Surface area*** |  |  |  |  |  |  |
| Frontal lobe | 11,902 | -0.007 | 0.008 | -0.860 | 0.392 | 0.756 |
| Temporal lobe | 11,904 | -0.014 | 0.008 | -1.700 | 0.088 | 0.380 |
| Occipital lobe | 11,898 | 0.007 | 0.011 | 0.570 | 0.567 | 0.756 |
| Parietal lobe | 11,900 | 0.003 | 0.009 | 0.280 | 0.781 | 0.893 |
| Cingulate lobe | 11,912 | -0.005 | 0.009 | -0.580 | 0.560 | 0.756 |
| Insula lobe | 11,912 | 0.001 | 0.010 | 0.110 | 0.909 | 0.909 |
| Postcentral lobe | 11,912 | 0.007 | 0.009 | 0.750 | 0.453 | 0.756 |
| Paracentral lobe | 11,912 | -0.017 | 0.010 | -1.670 | 0.095 | 0.380 |
| ***Cortical volume*** |  |  |  |  |  |  |
| Frontal lobe | 11,909 | 0.003 | 0.009 | 0.300 | 0.765 | 0.896 |
| Temporal lobe | 11,906 | -0.010 | 0.009 | -1.100 | 0.270 | 0.896 |
| Occipital lobe | 11,906 | 0.003 | 0.011 | 0.300 | 0.768 | 0.896 |
| Parietal lobe | 11,912 | 0.008 | 0.009 | 0.870 | 0.382 | 0.896 |
| Cingulate lobe | 11,912 | -0.007 | 0.010 | -0.720 | 0.473 | 0.896 |
| Insula lobe | 11,896 | -0.001 | 0.010 | -0.130 | 0.896 | 0.896 |
| Postcentral lobe | 11,910 | 0.002 | 0.010 | 0.170 | 0.863 | 0.896 |
| Paracentral lobe | 11,913 | -0.002 | 0.011 | -0.230 | 0.816 | 0.896 |
| ***Parcellations*** |  |  |  |  |  |  |
| ***Cortical thickness*** |  |  |  |  |  |  |
| Caudal anterior cingulate | 11,919 | 0.002 | 0.011 | 0.190 | 0.850 | 0.983 |
| Caudal middle frontal | 11,892 | 0.006 | 0.013 | 0.510 | 0.608 | 0.983 |
| Entorhinal | 11,901 | 0.009 | 0.012 | 0.720 | 0.470 | 0.983 |
| Fusiform | 11,913 | 0.005 | 0.013 | 0.430 | 0.665 | 0.983 |
| Inferior parietal | 11,889 | 0.001 | 0.013 | 0.060 | 0.953 | 0.983 |
| Inferior temporal | 11,917 | 0.014 | 0.013 | 1.080 | 0.279 | 0.983 |
| Isthmus cingulate | 11,917 | 0.004 | 0.012 | 0.290 | 0.769 | 0.983 |
| Lateral occipital | 11,910 | -0.008 | 0.013 | -0.640 | 0.520 | 0.983 |
| Lateral orbitofrontal | 11,911 | <0.001 | 0.013 | 0.020 | 0.983 | 0.983 |
| Medial orbitofrontal | 11,911 | 0.002 | 0.012 | 0.180 | 0.861 | 0.983 |
| Middle temporal | 11,918 | 0.013 | 0.013 | 1.030 | 0.304 | 0.983 |
| Parahippocampal | 11,916 | 0.008 | 0.013 | 0.620 | 0.533 | 0.983 |
| Paracentral | 11,912 | 0.018 | 0.013 | 1.410 | 0.158 | 0.983 |
| Postcentral | 11,918 | -0.003 | 0.013 | -0.260 | 0.797 | 0.983 |
| Posterior cingulate | 11,920 | 0.012 | 0.012 | 1.000 | 0.316 | 0.983 |
| Precentral | 11,899 | 0.005 | 0.013 | 0.410 | 0.683 | 0.983 |
| Precuneus | 11,896 | 0.009 | 0.013 | 0.700 | 0.481 | 0.983 |
| Rostral anterior cingulate | 11,917 | 0.006 | 0.011 | 0.540 | 0.587 | 0.983 |
| Superior parietal | 11,891 | <0.001 | 0.013 | -0.030 | 0.977 | 0.983 |
| Supramarginal | 11,887 | 0.003 | 0.013 | 0.250 | 0.799 | 0.983 |
| Frontal pole | 11,914 | 0.004 | 0.012 | 0.330 | 0.739 | 0.983 |
| Insula | 11,920 | 0.005 | 0.012 | 0.430 | 0.669 | 0.983 |
| Superior temporal | 11,906 | 0.006 | 0.013 | 0.470 | 0.635 | 0.983 |
| Inferior frontal | 11,891 | 0.005 | 0.013 | 0.410 | 0.683 | 0.983 |
| Dorsal lateral prefrontal | 11,875 | 0.026 | 0.013 | 2.030 | 0.042 | 0.983 |
| Medial occipital | 11,913 | -0.002 | 0.013 | -0.140 | 0.888 | 0.983 |
| ***Surface area*** |  |  |  |  |  |  |
| Caudal anterior cingulate | 11,914 | -0.014 | 0.010 | -1.440 | 0.149 | 0.719 |
| Caudal middle frontal | 11,910 | -0.014 | 0.010 | -1.300 | 0.194 | 0.719 |
| Entorhinal | 11,918 | -0.010 | 0.012 | -0.840 | 0.402 | 0.719 |
| Fusiform | 11,911 | 0.002 | 0.009 | 0.230 | 0.819 | 0.887 |
| Inferior parietal | 11,912 | -0.009 | 0.010 | -0.890 | 0.372 | 0.719 |
| Inferior temporal | 11,912 | -0.012 | 0.010 | -1.300 | 0.194 | 0.719 |
| Isthmus cingulate | 11,910 | 0.005 | 0.010 | 0.480 | 0.634 | 0.785 |
| Lateral occipital | 11,907 | -0.007 | 0.011 | -0.620 | 0.535 | 0.732 |
| Lateral orbitofrontal | 11,911 | 0.003 | 0.010 | 0.340 | 0.737 | 0.871 |
| Medial orbitofrontal | 11,912 | -0.009 | 0.009 | -1.060 | 0.289 | 0.719 |
| Middle temporal | 11,904 | -0.016 | 0.010 | -1.710 | 0.087 | 0.719 |
| Parahippocampal | 11,913 | 0.002 | 0.011 | 0.240 | 0.812 | 0.887 |
| Paracentral | 11,912 | -0.017 | 0.010 | -1.670 | 0.095 | 0.719 |
| Postcentral | 11,912 | 0.007 | 0.009 | 0.750 | 0.453 | 0.719 |
| Posterior cingulate | 11,910 | <0.001 | 0.010 | 0.030 | 0.973 | 0.973 |
| Precentral | 11,906 | -0.005 | 0.010 | -0.510 | 0.609 | 0.785 |
| Precuneus | 11,901 | 0.011 | 0.010 | 1.110 | 0.268 | 0.719 |
| Rostral anterior cingulate | 11,913 | -0.011 | 0.010 | -1.100 | 0.271 | 0.719 |
| Superior parietal | 11,904 | 0.009 | 0.011 | 0.840 | 0.399 | 0.719 |
| Supramarginal | 11,910 | -0.007 | 0.010 | -0.720 | 0.469 | 0.719 |
| Frontal pole | 11,916 | 0.007 | 0.010 | 0.680 | 0.498 | 0.719 |
| Insula | 11,912 | 0.001 | 0.010 | 0.110 | 0.909 | 0.945 |
| Superior temporal | 11,910 | -0.017 | 0.009 | -1.870 | 0.062 | 0.719 |
| Inferior frontal | 11,910 | -0.007 | 0.010 | -0.710 | 0.480 | 0.719 |
| Dorsal lateral prefrontal | 11,900 | -0.007 | 0.008 | -0.770 | 0.441 | 0.719 |
| Medial occipital | 11,907 | 0.016 | 0.012 | 1.330 | 0.184 | 0.719 |
| ***Cortical volume*** |  |  |  |  |  |  |
| Caudal anterior cingulate | 11,918 | -0.013 | 0.010 | -1.260 | 0.206 | 0.982 |
| Caudal middle frontal | 11,912 | -0.006 | 0.011 | -0.560 | 0.576 | 0.982 |
| Entorhinal | 11,912 | 0.001 | 0.012 | 0.070 | 0.945 | 0.982 |
| Fusiform | 11,914 | 0.005 | 0.010 | 0.460 | 0.646 | 0.982 |
| Inferior parietal | 11,914 | -0.002 | 0.010 | -0.190 | 0.848 | 0.982 |
| Inferior temporal | 11,914 | -0.010 | 0.010 | -1.040 | 0.296 | 0.982 |
| Isthmus cingulate | 11,915 | 0.009 | 0.010 | 0.920 | 0.359 | 0.982 |
| Lateral occipital | 11,912 | -0.007 | 0.010 | -0.680 | 0.494 | 0.982 |
| Lateral orbitofrontal | 11,905 | 0.001 | 0.010 | 0.060 | 0.955 | 0.982 |
| Medial orbitofrontal | 11,916 | -0.006 | 0.010 | -0.560 | 0.573 | 0.982 |
| Middle temporal | 11,910 | -0.008 | 0.010 | -0.800 | 0.424 | 0.982 |
| Parahippocampal | 11,916 | 0.005 | 0.012 | 0.420 | 0.673 | 0.982 |
| Paracentral | 11,913 | -0.002 | 0.011 | -0.230 | 0.816 | 0.982 |
| Postcentral | 11,910 | 0.002 | 0.010 | 0.170 | 0.863 | 0.982 |
| Posterior cingulate | 11,916 | -0.003 | 0.010 | -0.330 | 0.745 | 0.982 |
| Precentral | 11,912 | <0.001 | 0.010 | 0.020 | 0.982 | 0.982 |
| Precuneus | 11,903 | 0.018 | 0.010 | 1.800 | 0.071 | 0.982 |
| Rostral anterior cingulate | 11,917 | -0.010 | 0.010 | -1.000 | 0.316 | 0.982 |
| Superior parietal | 11,915 | 0.014 | 0.011 | 1.310 | 0.190 | 0.982 |
| Supramarginal | 11,911 | -0.004 | 0.010 | -0.360 | 0.721 | 0.982 |
| Frontal pole | 11,906 | 0.006 | 0.011 | 0.520 | 0.602 | 0.982 |
| Insula | 11,896 | -0.001 | 0.010 | -0.130 | 0.896 | 0.982 |
| Superior temporal | 11,901 | -0.015 | 0.010 | -1.490 | 0.137 | 0.982 |
| Inferior frontal | 11,906 | -0.004 | 0.011 | -0.420 | 0.675 | 0.982 |
| Dorsal lateral prefrontal | 11,909 | 0.006 | 0.009 | 0.730 | 0.466 | 0.982 |
| Medial occipital | 11,904 | 0.014 | 0.012 | 1.140 | 0.254 | 0.982 |
| ***Subcortical volumes*** |  |  |  |  |  |  |
| Thalamus | 11,894 | 0.005 | 0.009 | 0.490 | 0.623 | 0.850 |
| Caudate | 11,890 | -0.008 | 0.012 | -0.700 | 0.486 | 0.850 |
| Putamen | 11,881 | 0.007 | 0.011 | 0.630 | 0.529 | 0.850 |
| Pallidum | 11,904 | -0.001 | 0.011 | -0.140 | 0.892 | 0.892 |
| Hippocampus | 11,908 | -0.004 | 0.011 | -0.330 | 0.744 | 0.850 |
| Amygdala | 11,902 | -0.008 | 0.010 | -0.740 | 0.459 | 0.850 |
| Accumbens | 11,911 | -0.010 | 0.011 | -0.940 | 0.348 | 0.850 |
| Left hippocampus | 11,877 | -0.003 | 0.012 | -0.290 | 0.773 | 0.850 |
| Right hippocampus | 11,879 | -0.004 | 0.012 | -0.380 | 0.701 | 0.850 |
| Left accumbens | 11,873 | -0.008 | 0.012 | -0.620 | 0.536 | 0.850 |
| Right accumbens | 11,872 | -0.012 | 0.012 | -0.960 | 0.337 | 0.850 |

#### Table S9. The interaction between polygenic risk score for schizophrenia and Townsend deprivation index on brain structures.

| **Brain structure** | **N** | **β** | **SE** | **z** | **p_uncorrected_** | **p_corrected_** |
| --- | --- | --- | --- | --- | --- | --- |
| ***Global*** |  |  |  |  |  |  |
| Cortical thickness | 18,016 | -0.002 | 0.003 | -0.850 | 0.396 | 0.571 |
| Surface area | 18,037 | 0.001 | 0.001 | 0.610 | 0.542 | 0.571 |
| Cortical volume | 18,050 | 0.001 | 0.002 | 0.570 | 0.571 | 0.571 |
| ***Lobes*** |  |  |  |  |  |  |
| ***Cortical thickness*** |  |  |  |  |  |  |
| Frontal lobe | 18,019 | -0.001 | 0.003 | -0.330 | 0.741 | 0.969 |
| Temporal lobe | 18,044 | <0.001 | 0.002 | -0.040 | 0.969 | 0.969 |
| Occipital lobe | 18,061 | -0.001 | 0.003 | -0.520 | 0.600 | 0.960 |
| Parietal lobe | 17,989 | -0.002 | 0.003 | -0.910 | 0.365 | 0.960 |
| Cingulate lobe | 18,066 | 0.003 | 0.002 | 1.380 | 0.167 | 0.960 |
| Insula lobe | 18,069 | -0.003 | 0.002 | -1.120 | 0.262 | 0.960 |
| Postcentral lobe | 18,059 | <0.001 | 0.003 | 0.080 | 0.939 | 0.969 |
| Paracentral lobe | 18,058 | -0.002 | 0.003 | -0.590 | 0.553 | 0.960 |
| ***Surface area*** |  |  |  |  |  |  |
| Frontal lobe | 18,042 | <0.001 | 0.002 | -0.180 | 0.855 | 0.888 |
| Temporal lobe | 18,047 | -0.001 | 0.002 | -0.740 | 0.459 | 0.888 |
| Occipital lobe | 18,043 | 0.004 | 0.002 | 1.650 | 0.100 | 0.544 |
| Parietal lobe | 18,044 | 0.002 | 0.002 | 1.270 | 0.204 | 0.544 |
| Cingulate lobe | 18,058 | <0.001 | 0.002 | -0.140 | 0.888 | 0.888 |
| Insula lobe | 18,057 | 0.001 | 0.002 | 0.380 | 0.706 | 0.888 |
| Postcentral lobe | 18,057 | <0.001 | 0.002 | 0.140 | 0.886 | 0.888 |
| Paracentral lobe | 18,055 | 0.003 | 0.002 | 1.300 | 0.194 | 0.544 |
| ***Cortical volume*** |  |  |  |  |  |  |
| Frontal lobe | 18,048 | <0.001 | 0.002 | 0.210 | 0.831 | 0.856 |
| Temporal lobe | 18,052 | -0.001 | 0.002 | -0.770 | 0.439 | 0.731 |
| Occipital lobe | 18,053 | 0.003 | 0.002 | 1.190 | 0.232 | 0.712 |
| Parietal lobe | 18,058 | 0.002 | 0.002 | 1.110 | 0.267 | 0.712 |
| Cingulate lobe | 18,060 | 0.001 | 0.002 | 0.600 | 0.548 | 0.731 |
| Insula lobe | 18,038 | <0.001 | 0.002 | -0.180 | 0.856 | 0.856 |
| Postcentral lobe | 18,056 | 0.001 | 0.002 | 0.720 | 0.471 | 0.731 |
| Paracentral lobe | 18,062 | 0.003 | 0.002 | 1.200 | 0.230 | 0.712 |
| ***Parcellations*** |  |  |  |  |  |  |
| ***Cortical thickness*** |  |  |  |  |  |  |
| Caudal anterior cingulate | 18,069 | 0.003 | 0.002 | 1.590 | 0.112 | 0.943 |
| Caudal middle frontal | 18,022 | -0.001 | 0.002 | -0.560 | 0.577 | 0.943 |
| Entorhinal | 18,036 | -0.002 | 0.002 | -0.900 | 0.368 | 0.943 |
| Fusiform | 18,054 | <0.001 | 0.002 | -0.020 | 0.985 | 0.985 |
| Inferior parietal | 18,015 | -0.001 | 0.002 | -0.220 | 0.829 | 0.943 |
| Inferior temporal | 18,062 | -0.001 | 0.002 | -0.430 | 0.664 | 0.943 |
| Isthmus cingulate | 18,066 | <0.001 | 0.002 | -0.160 | 0.870 | 0.943 |
| Lateral occipital | 18,058 | -0.001 | 0.003 | -0.360 | 0.721 | 0.943 |
| Lateral orbitofrontal | 18,055 | -0.002 | 0.003 | -0.840 | 0.404 | 0.943 |
| Medial orbitofrontal | 18,057 | 0.001 | 0.002 | 0.410 | 0.684 | 0.943 |
| Middle temporal | 18,066 | -0.001 | 0.002 | -0.250 | 0.805 | 0.943 |
| Parahippocampal | 18,062 | 0.005 | 0.002 | 1.870 | 0.062 | 0.943 |
| Paracentral | 18,058 | -0.002 | 0.003 | -0.590 | 0.553 | 0.943 |
| Postcentral | 18,059 | <0.001 | 0.003 | 0.080 | 0.939 | 0.977 |
| Posterior cingulate | 18,068 | 0.003 | 0.002 | 1.090 | 0.274 | 0.943 |
| Precentral | 18,036 | <0.001 | 0.002 | -0.170 | 0.863 | 0.943 |
| Precuneus | 18,031 | -0.002 | 0.003 | -0.650 | 0.516 | 0.943 |
| Rostral anterior cingulate | 18,065 | 0.001 | 0.002 | 0.620 | 0.538 | 0.943 |
| Superior parietal | 18,017 | -0.001 | 0.003 | -0.240 | 0.813 | 0.943 |
| Supramarginal | 18,015 | -0.003 | 0.002 | -1.340 | 0.181 | 0.943 |
| Frontal pole | 18,058 | 0.002 | 0.002 | 0.830 | 0.408 | 0.943 |
| Insula | 18,069 | -0.003 | 0.002 | -1.120 | 0.262 | 0.943 |
| Superior temporal | 18,041 | -0.003 | 0.003 | -1.100 | 0.273 | 0.943 |
| Inferior frontal | 18,021 | -0.003 | 0.002 | -1.130 | 0.259 | 0.943 |
| Dorsal lateral prefrontal | 17,984 | -0.001 | 0.002 | -0.500 | 0.618 | 0.943 |
| Medial occipital | 18,059 | -0.002 | 0.003 | -0.660 | 0.507 | 0.943 |
| ***Surface area*** |  |  |  |  |  |  |
| Caudal anterior cingulate | 18,061 | -0.001 | 0.002 | -0.280 | 0.778 | 0.902 |
| Caudal middle frontal | 18,051 | <0.001 | 0.002 | -0.110 | 0.916 | 0.916 |
| Entorhinal | 18,065 | 0.002 | 0.002 | 0.940 | 0.345 | 0.637 |
| Fusiform | 18,059 | 0.001 | 0.002 | 0.660 | 0.507 | 0.739 |
| Inferior parietal | 18,062 | 0.001 | 0.002 | 0.260 | 0.798 | 0.902 |
| Inferior temporal | 18,061 | -0.003 | 0.002 | -1.460 | 0.145 | 0.624 |
| Isthmus cingulate | 18,052 | -0.001 | 0.002 | -0.610 | 0.540 | 0.739 |
| Lateral occipital | 18,054 | 0.003 | 0.002 | 1.240 | 0.214 | 0.624 |
| Lateral orbitofrontal | 18,059 | 0.002 | 0.002 | 0.880 | 0.377 | 0.637 |
| Medial orbitofrontal | 18,058 | 0.002 | 0.002 | 0.920 | 0.357 | 0.637 |
| Middle temporal | 18,051 | -0.003 | 0.002 | -1.600 | 0.110 | 0.624 |
| Parahippocampal | 18,060 | 0.003 | 0.002 | 1.350 | 0.177 | 0.624 |
| Paracentral | 18,055 | 0.003 | 0.002 | 1.300 | 0.194 | 0.624 |
| Postcentral | 18,057 | <0.001 | 0.002 | 0.140 | 0.886 | 0.916 |
| Posterior cingulate | 18,053 | 0.002 | 0.002 | 1.130 | 0.259 | 0.624 |
| Precentral | 18,055 | 0.002 | 0.002 | 0.860 | 0.392 | 0.637 |
| Precuneus | 18,040 | 0.003 | 0.002 | 1.300 | 0.195 | 0.624 |
| Rostral anterior cingulate | 18,060 | -0.002 | 0.002 | -1.010 | 0.314 | 0.637 |
| Superior parietal | 18,049 | 0.003 | 0.002 | 1.240 | 0.214 | 0.624 |
| Supramarginal | 18,056 | <0.001 | 0.002 | -0.120 | 0.904 | 0.916 |
| Frontal pole | 18,067 | 0.001 | 0.002 | 0.510 | 0.607 | 0.789 |
| Insula | 18,057 | 0.001 | 0.002 | 0.380 | 0.706 | 0.874 |
| Superior temporal | 18,057 | -0.002 | 0.002 | -1.220 | 0.223 | 0.624 |
| Inferior frontal | 18,050 | -0.002 | 0.002 | -1.120 | 0.264 | 0.624 |
| Dorsal lateral prefrontal | 18,041 | -0.001 | 0.002 | -0.620 | 0.535 | 0.739 |
| Medial occipital | 18,050 | 0.004 | 0.002 | 1.490 | 0.136 | 0.624 |
| ***Cortical volume*** |  |  |  |  |  |  |
| Caudal anterior cingulate | 18,069 | 0.002 | 0.002 | 0.880 | 0.378 | 0.630 |
| Caudal middle frontal | 18,055 | 0.001 | 0.002 | 0.410 | 0.682 | 0.771 |
| Entorhinal | 18,058 | <0.001 | 0.002 | 0.210 | 0.835 | 0.856 |
| Fusiform | 18,062 | 0.002 | 0.002 | 0.760 | 0.444 | 0.630 |
| Inferior parietal | 18,061 | 0.001 | 0.002 | 0.660 | 0.509 | 0.630 |
| Inferior temporal | 18,062 | -0.002 | 0.002 | -0.930 | 0.352 | 0.630 |
| Isthmus cingulate | 18,062 | -0.002 | 0.002 | -1.020 | 0.306 | 0.630 |
| Lateral occipital | 18,058 | 0.003 | 0.002 | 1.370 | 0.170 | 0.553 |
| Lateral orbitofrontal | 18,047 | 0.001 | 0.002 | 0.670 | 0.501 | 0.630 |
| Medial orbitofrontal | 18,064 | 0.004 | 0.002 | 1.880 | 0.060 | 0.553 |
| Middle temporal | 18,056 | -0.003 | 0.002 | -1.720 | 0.085 | 0.553 |
| Parahippocampal | 18,065 | 0.006 | 0.002 | 2.410 | 0.016 | 0.416 |
| Paracentral | 18,062 | 0.003 | 0.002 | 1.200 | 0.230 | 0.630 |
| Postcentral | 18,056 | 0.001 | 0.002 | 0.720 | 0.471 | 0.630 |
| Posterior cingulate | 18,063 | 0.004 | 0.002 | 1.800 | 0.072 | 0.553 |
| Precentral | 18,056 | 0.001 | 0.002 | 0.710 | 0.478 | 0.630 |
| Precuneus | 18,038 | 0.003 | 0.002 | 1.400 | 0.163 | 0.553 |
| Rostral anterior cingulate | 18,067 | -0.002 | 0.002 | -0.940 | 0.347 | 0.630 |
| Superior parietal | 18,062 | 0.003 | 0.002 | 1.380 | 0.167 | 0.553 |
| Supramarginal | 18,057 | -0.001 | 0.002 | -0.530 | 0.594 | 0.702 |
| Frontal pole | 18,044 | 0.003 | 0.002 | 1.520 | 0.130 | 0.553 |
| Insula | 18,038 | <0.001 | 0.002 | -0.180 | 0.856 | 0.856 |
| Superior temporal | 18,045 | -0.002 | 0.002 | -0.960 | 0.338 | 0.630 |
| Inferior frontal | 18,051 | -0.002 | 0.002 | -0.870 | 0.387 | 0.630 |
| Dorsal lateral prefrontal | 18,051 | <0.001 | 0.002 | -0.210 | 0.833 | 0.856 |
| Medial occipital | 18,049 | 0.002 | 0.002 | 0.790 | 0.429 | 0.630 |
| ***Subcortical volumes*** |  |  |  |  |  |  |
| Thalamus | 18,037 | 0.003 | 0.002 | 1.520 | 0.128 | 0.660 |
| Caudate | 18,009 | -0.004 | 0.002 | -1.820 | 0.069 | 0.660 |
| Putamen | 18,012 | 0.001 | 0.002 | 0.500 | 0.619 | 0.757 |
| Pallidum | 18,044 | -0.002 | 0.002 | -0.770 | 0.442 | 0.757 |
| Hippocampus | 18,047 | 0.002 | 0.002 | 0.940 | 0.345 | 0.757 |
| Amygdala | 18,049 | 0.003 | 0.002 | 1.340 | 0.180 | 0.660 |
| Accumbens | 18,049 | <0.001 | 0.002 | 0.220 | 0.823 | 0.828 |
| Left hippocampus | 17,997 | 0.003 | 0.002 | 1.170 | 0.243 | 0.668 |
| Right hippocampus | 17,995 | 0.001 | 0.002 | 0.610 | 0.540 | 0.757 |
| Left accumbens | 17,985 | 0.001 | 0.002 | 0.580 | 0.560 | 0.757 |
| Right accumbens | 17,988 | -0.001 | 0.002 | -0.220 | 0.828 | 0.828 |

#### Table S10. Main effect of childhood trauma on brain structures in the interaction model.

| **Brain structure** | **N** | **β** | **SE** | **z** | **p_uncorrected_** | **p_corrected_** |
| --- | --- | --- | --- | --- | --- | --- |
| ***Global*** |  |  |  |  |  |  |
| Cortical thickness | 12,963 | -0.003 | 0.003 | -0.950 | 0.345 | 0.345 |
| Surface area | 12,985 | -0.004 | 0.002 | -2.390 | 0.017 | 0.026 |
| Cortical volume | 12,990 | -0.005 | 0.002 | -2.640 | 0.008 | 0.024 |
| ***Lobes*** |  |  |  |  |  |  |
| ***Cortical thickness*** |  |  |  |  |  |  |
| Frontal lobe | 12,965 | -0.005 | 0.003 | -1.580 | 0.115 | 0.526 |
| Temporal lobe | 12,980 | -0.002 | 0.003 | -0.620 | 0.535 | 0.856 |
| Occipital lobe | 12,994 | 0.004 | 0.003 | 1.250 | 0.212 | 0.526 |
| Parietal lobe | 12,947 | -0.004 | 0.003 | -1.240 | 0.216 | 0.526 |
| Cingulate lobe | 12,997 | <0.001 | 0.003 | 0.010 | 0.996 | 0.996 |
| Insula lobe | 12,999 | <0.001 | 0.003 | -0.050 | 0.957 | 0.996 |
| Postcentral lobe | 12,992 | <0.001 | 0.003 | -0.100 | 0.921 | 0.996 |
| Paracentral lobe | 12,990 | -0.004 | 0.003 | -1.120 | 0.263 | 0.526 |
| ***Surface area*** |  |  |  |  |  |  |
| Frontal lobe | 12,984 | -0.002 | 0.002 | -0.900 | 0.370 | 0.583 |
| Temporal lobe | 12,985 | -0.006 | 0.002 | -2.670 | 0.008 | 0.064 |
| Occipital lobe | 12,984 | -0.006 | 0.003 | -2.170 | 0.030 | 0.080 |
| Parietal lobe | 12,989 | -0.005 | 0.002 | -2.230 | 0.026 | 0.080 |
| Cingulate lobe | 12,991 | 0.002 | 0.002 | 0.780 | 0.437 | 0.583 |
| Insula lobe | 12,993 | -0.001 | 0.002 | -0.420 | 0.672 | 0.768 |
| Postcentral lobe | 12,990 | -0.005 | 0.002 | -2.000 | 0.046 | 0.092 |
| Paracentral lobe | 12,990 | <0.001 | 0.003 | -0.110 | 0.913 | 0.913 |
| ***Cortical volume*** |  |  |  |  |  |  |
| Frontal lobe | 12,989 | -0.005 | 0.002 | -2.190 | 0.028 | 0.075 |
| Temporal lobe | 12,989 | -0.006 | 0.002 | -2.490 | 0.013 | 0.052 |
| Occipital lobe | 12,991 | -0.002 | 0.003 | -0.820 | 0.410 | 0.547 |
| Parietal lobe | 12,992 | -0.007 | 0.002 | -3.020 | 0.003 | 0.024 |
| Cingulate lobe | 12,993 | 0.001 | 0.002 | 0.570 | 0.568 | 0.568 |
| Insula lobe | 12,979 | -0.002 | 0.003 | -0.640 | 0.522 | 0.568 |
| Postcentral lobe | 12,994 | -0.004 | 0.003 | -1.390 | 0.165 | 0.330 |
| Paracentral lobe | 12,996 | -0.003 | 0.003 | -1.160 | 0.247 | 0.395 |
| ***Parcellations*** |  |  |  |  |  |  |
| ***Cortical thickness*** |  |  |  |  |  |  |
| Caudal anterior cingulate | 12,999 | 0.001 | 0.003 | 0.230 | 0.820 | 0.927 |
| Caudal middle frontal | 12,966 | -0.003 | 0.003 | -1.040 | 0.297 | 0.594 |
| Entorhinal | 12,974 | -0.001 | 0.003 | -0.480 | 0.630 | 0.780 |
| Fusiform | 12,986 | -0.003 | 0.003 | -0.830 | 0.405 | 0.660 |
| Inferior parietal | 12,961 | -0.007 | 0.003 | -2.250 | 0.025 | 0.321 |
| Inferior temporal | 12,995 | -0.006 | 0.003 | -1.850 | 0.064 | 0.333 |
| Isthmus cingulate | 12,997 | 0.004 | 0.003 | 1.240 | 0.216 | 0.570 |
| Lateral occipital | 12,992 | 0.004 | 0.003 | 1.160 | 0.248 | 0.570 |
| Lateral orbitofrontal | 12,988 | -0.002 | 0.003 | -0.740 | 0.457 | 0.699 |
| Medial orbitofrontal | 12,989 | -0.008 | 0.003 | -2.470 | 0.013 | 0.321 |
| Middle temporal | 12,998 | -0.005 | 0.003 | -1.470 | 0.141 | 0.533 |
| Parahippocampal | 12,995 | <0.001 | 0.003 | 0.150 | 0.877 | 0.950 |
| Paracentral | 12,990 | -0.004 | 0.003 | -1.120 | 0.263 | 0.570 |
| Postcentral | 12,992 | <0.001 | 0.003 | -0.100 | 0.921 | 0.957 |
| Posterior cingulate | 12,999 | -0.002 | 0.003 | -0.610 | 0.545 | 0.749 |
| Precentral | 12,975 | -0.007 | 0.003 | -2.080 | 0.037 | 0.321 |
| Precuneus | 12,977 | -0.003 | 0.003 | -0.830 | 0.406 | 0.660 |
| Rostral anterior cingulate | 12,995 | -0.002 | 0.003 | -0.830 | 0.404 | 0.660 |
| Superior parietal | 12,966 | -0.002 | 0.003 | -0.600 | 0.547 | 0.749 |
| Supramarginal | 12,964 | -0.005 | 0.003 | -1.450 | 0.146 | 0.533 |
| Frontal pole | 12,993 | -0.002 | 0.003 | -0.540 | 0.589 | 0.766 |
| Insula | 12,999 | <0.001 | 0.003 | -0.050 | 0.957 | 0.957 |
| Superior temporal | 12,978 | 0.001 | 0.003 | 0.310 | 0.759 | 0.897 |
| Inferior frontal | 12,964 | -0.006 | 0.003 | -1.870 | 0.062 | 0.333 |
| Dorsal lateral prefrontal | 12,940 | -0.005 | 0.003 | -1.390 | 0.164 | 0.533 |
| Medial occipital | 12,990 | 0.004 | 0.003 | 1.160 | 0.244 | 0.570 |
| ***Surface area*** |  |  |  |  |  |  |
| Caudal anterior cingulate | 12,993 | 0.001 | 0.002 | 0.320 | 0.750 | 0.813 |
| Caudal middle frontal | 12,989 | 0.004 | 0.003 | 1.350 | 0.178 | 0.356 |
| Entorhinal | 13,000 | -0.002 | 0.003 | -0.680 | 0.496 | 0.679 |
| Fusiform | 12,991 | -0.004 | 0.002 | -1.610 | 0.108 | 0.255 |
| Inferior parietal | 12,996 | -0.007 | 0.003 | -2.800 | 0.005 | 0.130 |
| Inferior temporal | 12,992 | -0.005 | 0.002 | -2.130 | 0.033 | 0.215 |
| Isthmus cingulate | 12,988 | 0.001 | 0.003 | 0.490 | 0.625 | 0.760 |
| Lateral occipital | 12,988 | -0.004 | 0.003 | -1.670 | 0.095 | 0.255 |
| Lateral orbitofrontal | 12,993 | -0.001 | 0.002 | -0.430 | 0.664 | 0.760 |
| Medial orbitofrontal | 12,992 | -0.002 | 0.002 | -0.980 | 0.326 | 0.557 |
| Middle temporal | 12,985 | -0.004 | 0.002 | -1.730 | 0.084 | 0.255 |
| Parahippocampal | 12,994 | -0.007 | 0.003 | -2.480 | 0.013 | 0.147 |
| Paracentral | 12,990 | <0.001 | 0.003 | -0.110 | 0.913 | 0.938 |
| Postcentral | 12,990 | -0.005 | 0.002 | -2.000 | 0.046 | 0.239 |
| Posterior cingulate | 12,988 | 0.002 | 0.002 | 0.690 | 0.493 | 0.679 |
| Precentral | 12,992 | -0.004 | 0.002 | -1.480 | 0.138 | 0.299 |
| Precuneus | 12,982 | -0.002 | 0.003 | -0.950 | 0.343 | 0.557 |
| Rostral anterior cingulate | 12,991 | 0.001 | 0.002 | 0.530 | 0.598 | 0.760 |
| Superior parietal | 12,986 | -0.002 | 0.003 | -0.900 | 0.367 | 0.561 |
| Supramarginal | 12,991 | -0.004 | 0.002 | -1.630 | 0.103 | 0.255 |
| Frontal pole | 12,997 | -0.005 | 0.003 | -1.900 | 0.057 | 0.247 |
| Insula | 12,993 | -0.001 | 0.002 | -0.420 | 0.672 | 0.760 |
| Superior temporal | 12,991 | -0.004 | 0.002 | -1.620 | 0.106 | 0.255 |
| Inferior frontal | 12,989 | <0.001 | 0.003 | 0.080 | 0.938 | 0.938 |
| Dorsal lateral prefrontal | 12,983 | -0.002 | 0.002 | -1.110 | 0.269 | 0.500 |
| Medial occipital | 12,986 | -0.007 | 0.003 | -2.380 | 0.017 | 0.147 |
| ***Cortical volume*** |  |  |  |  |  |  |
| Caudal anterior cingulate | 13,000 | 0.001 | 0.003 | 0.340 | 0.732 | 0.827 |
| Caudal middle frontal | 12,992 | 0.002 | 0.003 | 0.670 | 0.501 | 0.646 |
| Entorhinal | 12,995 | -0.001 | 0.003 | -0.360 | 0.716 | 0.827 |
| Fusiform | 12,995 | -0.003 | 0.003 | -1.280 | 0.199 | 0.370 |
| Inferior parietal | 12,995 | -0.009 | 0.003 | -3.580 | <0.001 | <0.001 |
| Inferior temporal | 12,995 | -0.006 | 0.002 | -2.370 | 0.018 | 0.099 |
| Isthmus cingulate | 12,994 | 0.003 | 0.003 | 1.230 | 0.219 | 0.380 |
| Lateral occipital | 12,992 | -0.001 | 0.003 | -0.300 | 0.765 | 0.829 |
| Lateral orbitofrontal | 12,988 | -0.002 | 0.003 | -0.890 | 0.374 | 0.540 |
| Medial orbitofrontal | 12,996 | -0.007 | 0.003 | -2.650 | 0.008 | 0.099 |
| Middle temporal | 12,990 | -0.006 | 0.002 | -2.350 | 0.019 | 0.099 |
| Parahippocampal | 12,997 | -0.005 | 0.003 | -1.720 | 0.086 | 0.248 |
| Paracentral | 12,996 | -0.003 | 0.003 | -1.160 | 0.247 | 0.401 |
| Postcentral | 12,994 | -0.004 | 0.003 | -1.390 | 0.165 | 0.332 |
| Posterior cingulate | 12,994 | <0.001 | 0.003 | -0.130 | 0.898 | 0.898 |
| Precentral | 12,989 | -0.007 | 0.003 | -2.470 | 0.014 | 0.099 |
| Precuneus | 12,979 | -0.004 | 0.003 | -1.440 | 0.150 | 0.332 |
| Rostral anterior cingulate | 12,998 | <0.001 | 0.003 | -0.170 | 0.867 | 0.898 |
| Superior parietal | 12,995 | -0.004 | 0.003 | -1.380 | 0.166 | 0.332 |
| Supramarginal | 12,995 | -0.005 | 0.003 | -1.860 | 0.063 | 0.224 |
| Frontal pole | 12,988 | -0.004 | 0.003 | -1.560 | 0.118 | 0.307 |
| Insula | 12,979 | -0.002 | 0.003 | -0.640 | 0.522 | 0.646 |
| Superior temporal | 12,980 | -0.005 | 0.003 | -1.820 | 0.069 | 0.224 |
| Inferior frontal | 12,987 | -0.002 | 0.003 | -0.740 | 0.459 | 0.628 |
| Dorsal lateral prefrontal | 12,989 | -0.005 | 0.002 | -2.210 | 0.027 | 0.117 |
| Medial occipital | 12,988 | -0.003 | 0.003 | -0.980 | 0.328 | 0.502 |
| ***Subcortical volumes*** |  |  |  |  |  |  |
| Thalamus | 12,972 | -0.004 | 0.002 | -1.860 | 0.063 | 0.347 |
| Caudate | 12,960 | 0.001 | 0.003 | 0.490 | 0.625 | 0.791 |
| Putamen | 12,958 | 0.003 | 0.003 | 1.160 | 0.245 | 0.791 |
| Pallidum | 12,981 | -0.007 | 0.003 | -2.510 | 0.012 | 0.132 |
| Hippocampus | 12,984 | -0.001 | 0.003 | -0.470 | 0.639 | 0.791 |
| Amygdala | 12,983 | 0.001 | 0.003 | 0.560 | 0.573 | 0.791 |
| Accumbens | 12,985 | -0.001 | 0.003 | -0.300 | 0.768 | 0.845 |
| Left hippocampus | 12,952 | -0.001 | 0.003 | -0.500 | 0.618 | 0.791 |
| Right hippocampus | 12,949 | -0.001 | 0.003 | -0.460 | 0.647 | 0.791 |
| Left accumbens | 12,940 | <0.001 | 0.003 | 0.100 | 0.920 | 0.920 |
| Right accumbens | 12,942 | -0.002 | 0.003 | -0.550 | 0.581 | 0.791 |

#### Table S11. Main effect of cannabis use on brain structures in the interaction model.

| **Brain structure** | **N** | **β** | **SE** | **z** | **p_uncorrected_** | **p_corrected_** |
| --- | --- | --- | --- | --- | --- | --- |
| ***Global*** |  |  |  |  |  |  |
| Cortical thickness | 13,215 | -0.043 | 0.019 | -2.250 | 0.025 | 0.038 |
| Surface area | 13,237 | 0.027 | 0.011 | 2.500 | 0.012 | 0.036 |
| Cortical volume | 13,242 | <0.001 | 0.012 | 0.040 | 0.970 | 0.970 |
| ***Lobes*** |  |  |  |  |  |  |
| ***Cortical thickness*** | |  |  |  |  |  |
| Frontal lobe | 13,217 | -0.048 | 0.019 | -2.530 | 0.011 | 0.044 |
| Temporal lobe | 13,230 | -0.021 | 0.019 | -1.150 | 0.249 | 0.398 |
| Occipital lobe | 13,246 | -0.030 | 0.020 | -1.520 | 0.129 | 0.344 |
| Parietal lobe | 13,198 | -0.062 | 0.019 | -3.240 | 0.001 | 0.008 |
| Cingulate lobe | 13,249 | -0.021 | 0.017 | -1.230 | 0.220 | 0.398 |
| Insula lobe | 13,251 | -0.005 | 0.018 | -0.280 | 0.783 | 0.783 |
| Postcentral lobe | 13,244 | -0.014 | 0.019 | -0.720 | 0.470 | 0.537 |
| Paracentral lobe | 13,242 | -0.016 | 0.019 | -0.830 | 0.404 | 0.537 |
| ***Surface area*** | |  |  |  |  |  |
| Frontal lobe | 13,236 | 0.021 | 0.012 | 1.790 | 0.074 | 0.118 |
| Temporal lobe | 13,236 | 0.035 | 0.012 | 2.920 | 0.004 | 0.016 |
| Occipital lobe | 13,235 | -0.005 | 0.017 | -0.310 | 0.755 | 0.755 |
| Parietal lobe | 13,241 | 0.022 | 0.013 | 1.670 | 0.095 | 0.127 |
| Cingulate lobe | 13,243 | 0.029 | 0.013 | 2.230 | 0.026 | 0.069 |
| Insula lobe | 13,245 | 0.042 | 0.014 | 2.930 | 0.003 | 0.016 |
| Postcentral lobe | 13,242 | 0.010 | 0.014 | 0.760 | 0.446 | 0.510 |
| Paracentral lobe | 13,242 | 0.031 | 0.015 | 2.090 | 0.037 | 0.074 |
| ***Cortical volume*** | |  |  |  |  |  |
| Frontal lobe | 13,241 | -0.004 | 0.013 | -0.280 | 0.781 | 0.811 |
| Temporal lobe | 13,241 | 0.010 | 0.013 | 0.730 | 0.464 | 0.742 |
| Occipital lobe | 13,243 | -0.004 | 0.016 | -0.240 | 0.811 | 0.811 |
| Parietal lobe | 13,244 | -0.011 | 0.014 | -0.770 | 0.440 | 0.742 |
| Cingulate lobe | 13,245 | 0.016 | 0.014 | 1.140 | 0.252 | 0.672 |
| Insula lobe | 13,231 | 0.037 | 0.015 | 2.450 | 0.014 | 0.112 |
| Postcentral lobe | 13,246 | 0.006 | 0.015 | 0.380 | 0.701 | 0.811 |
| Paracentral lobe | 13,248 | 0.027 | 0.016 | 1.730 | 0.084 | 0.336 |
| ***Parcellations*** |  |  |  |  |  |  |
| ***Cortical thickness*** | |  |  |  |  |  |
| Caudal anterior cingulate | 13,251 | -0.012 | 0.016 | -0.750 | 0.453 | 0.572 |
| Caudal middle frontal | 13,218 | -0.050 | 0.019 | -2.660 | 0.008 | 0.026 |
| Entorhinal | 13,225 | -0.018 | 0.018 | -1.010 | 0.312 | 0.477 |
| Fusiform | 13,238 | -0.044 | 0.019 | -2.390 | 0.017 | 0.040 |
| Inferior parietal | 13,213 | -0.060 | 0.019 | -3.250 | 0.001 | 0.007 |
| Inferior temporal | 13,247 | -0.057 | 0.018 | -3.110 | 0.002 | 0.010 |
| Isthmus cingulate | 13,249 | 0.029 | 0.018 | 1.580 | 0.115 | 0.221 |
| Lateral occipital | 13,244 | -0.047 | 0.019 | -2.420 | 0.015 | 0.040 |
| Lateral orbitofrontal | 13,240 | 0.001 | 0.019 | 0.060 | 0.950 | 0.950 |
| Medial orbitofrontal | 13,241 | -0.027 | 0.018 | -1.450 | 0.148 | 0.257 |
| Middle temporal | 13,250 | -0.024 | 0.018 | -1.280 | 0.200 | 0.325 |
| Parahippocampal | 13,246 | 0.012 | 0.019 | 0.630 | 0.527 | 0.596 |
| Paracentral | 13,242 | -0.016 | 0.019 | -0.830 | 0.404 | 0.572 |
| Postcentral | 13,244 | -0.014 | 0.019 | -0.720 | 0.470 | 0.572 |
| Posterior cingulate | 13,251 | -0.036 | 0.018 | -2.030 | 0.042 | 0.091 |
| Precentral | 13,226 | -0.013 | 0.019 | -0.710 | 0.477 | 0.572 |
| Precuneus | 13,229 | -0.052 | 0.019 | -2.690 | 0.007 | 0.026 |
| Rostral anterior cingulate | 13,247 | -0.026 | 0.017 | -1.560 | 0.119 | 0.221 |
| Superior parietal | 13,218 | -0.056 | 0.019 | -2.910 | 0.004 | 0.017 |
| Supramarginal | 13,216 | -0.061 | 0.019 | -3.290 | 0.001 | 0.007 |
| Frontal pole | 13,245 | -0.063 | 0.017 | -3.650 | <0.001 | <0.001 |
| Insula | 13,251 | -0.005 | 0.018 | -0.280 | 0.783 | 0.848 |
| Superior temporal | 13,230 | -0.013 | 0.019 | -0.700 | 0.484 | 0.572 |
| Inferior frontal | 13,216 | -0.045 | 0.019 | -2.410 | 0.016 | 0.040 |
| Dorsal lateral prefrontal | 13,190 | -0.081 | 0.019 | -4.300 | <0.001 | <0.001 |
| Medial occipital | 13,242 | -0.004 | 0.019 | -0.230 | 0.820 | 0.853 |
| ***Surface area*** | |  |  |  |  |  |
| Caudal anterior cingulate | 13,245 | 0.033 | 0.014 | 2.290 | 0.022 | 0.082 |
| Caudal middle frontal | 13,241 | 0.001 | 0.015 | 0.050 | 0.956 | 0.956 |
| Entorhinal | 13,252 | 0.063 | 0.017 | 3.580 | <0.001 | <0.001 |
| Fusiform | 13,243 | 0.039 | 0.014 | 2.810 | 0.005 | 0.033 |
| Inferior parietal | 13,248 | 0.022 | 0.015 | 1.480 | 0.138 | 0.294 |
| Inferior temporal | 13,244 | 0.027 | 0.014 | 1.940 | 0.053 | 0.138 |
| Isthmus cingulate | 13,240 | -0.014 | 0.015 | -0.980 | 0.326 | 0.424 |
| Lateral occipital | 13,240 | 0.018 | 0.016 | 1.130 | 0.257 | 0.371 |
| Lateral orbitofrontal | 13,245 | 0.002 | 0.014 | 0.160 | 0.870 | 0.905 |
| Medial orbitofrontal | 13,244 | 0.017 | 0.013 | 1.360 | 0.173 | 0.300 |
| Middle temporal | 13,237 | 0.036 | 0.014 | 2.570 | 0.010 | 0.052 |
| Parahippocampal | 13,246 | -0.007 | 0.015 | -0.460 | 0.647 | 0.765 |
| Paracentral | 13,242 | 0.031 | 0.015 | 2.090 | 0.037 | 0.120 |
| Postcentral | 13,242 | 0.010 | 0.014 | 0.760 | 0.446 | 0.552 |
| Posterior cingulate | 13,240 | 0.014 | 0.014 | 0.990 | 0.322 | 0.424 |
| Precentral | 13,244 | 0.005 | 0.014 | 0.360 | 0.718 | 0.802 |
| Precuneus | 13,233 | 0.005 | 0.015 | 0.330 | 0.740 | 0.802 |
| Rostral anterior cingulate | 13,242 | 0.053 | 0.014 | 3.700 | <0.001 | <0.001 |
| Superior parietal | 13,238 | 0.022 | 0.016 | 1.390 | 0.165 | 0.300 |
| Supramarginal | 13,242 | 0.021 | 0.014 | 1.450 | 0.147 | 0.294 |
| Frontal pole | 13,249 | 0.017 | 0.015 | 1.170 | 0.240 | 0.367 |
| Insula | 13,245 | 0.042 | 0.014 | 2.930 | 0.003 | 0.026 |
| Superior temporal | 13,242 | 0.025 | 0.014 | 1.850 | 0.064 | 0.151 |
| Inferior frontal | 13,241 | 0.030 | 0.015 | 2.000 | 0.046 | 0.133 |
| Dorsal lateral prefrontal | 13,235 | 0.029 | 0.013 | 2.290 | 0.022 | 0.082 |
| Medial occipital | 13,238 | -0.022 | 0.018 | -1.230 | 0.220 | 0.358 |
| ***Cortical volume*** | |  |  |  |  |  |
| Caudal anterior cingulate | 13,252 | 0.017 | 0.015 | 1.150 | 0.251 | 0.823 |
| Caudal middle frontal | 13,244 | -0.016 | 0.015 | -1.070 | 0.285 | 0.823 |
| Entorhinal | 13,247 | 0.036 | 0.017 | 2.120 | 0.034 | 0.295 |
| Fusiform | 13,247 | 0.021 | 0.015 | 1.410 | 0.159 | 0.689 |
| Inferior parietal | 13,247 | -0.005 | 0.015 | -0.330 | 0.740 | 0.960 |
| Inferior temporal | 13,246 | -0.006 | 0.014 | -0.420 | 0.673 | 0.960 |
| Isthmus cingulate | 13,246 | -0.006 | 0.015 | -0.360 | 0.719 | 0.960 |
| Lateral occipital | 13,244 | <0.001 | 0.015 | -0.010 | 0.992 | 0.992 |
| Lateral orbitofrontal | 13,240 | 0.010 | 0.015 | 0.690 | 0.493 | 0.960 |
| Medial orbitofrontal | 13,248 | -0.016 | 0.015 | -1.070 | 0.283 | 0.823 |
| Middle temporal | 13,242 | 0.012 | 0.014 | 0.860 | 0.391 | 0.847 |
| Parahippocampal | 13,249 | -0.003 | 0.018 | -0.140 | 0.886 | 0.960 |
| Paracentral | 13,248 | 0.027 | 0.016 | 1.730 | 0.084 | 0.546 |
| Postcentral | 13,246 | 0.006 | 0.015 | 0.380 | 0.701 | 0.960 |
| Posterior cingulate | 13,246 | <0.001 | 0.015 | 0.020 | 0.986 | 0.992 |
| Precentral | 13,241 | 0.003 | 0.015 | 0.160 | 0.871 | 0.960 |
| Precuneus | 13,231 | -0.014 | 0.015 | -0.970 | 0.333 | 0.847 |
| Rostral anterior cingulate | 13,250 | 0.035 | 0.015 | 2.350 | 0.019 | 0.247 |
| Superior parietal | 13,247 | -0.007 | 0.016 | -0.430 | 0.666 | 0.960 |
| Supramarginal | 13,246 | -0.003 | 0.015 | -0.220 | 0.824 | 0.960 |
| Frontal pole | 13,239 | -0.026 | 0.016 | -1.560 | 0.120 | 0.624 |
| Insula | 13,231 | 0.037 | 0.015 | 2.450 | 0.014 | 0.247 |
| Superior temporal | 13,232 | 0.007 | 0.015 | 0.440 | 0.660 | 0.960 |
| Inferior frontal | 13,239 | 0.014 | 0.015 | 0.880 | 0.379 | 0.847 |
| Dorsal lateral prefrontal | 13,241 | -0.008 | 0.013 | -0.590 | 0.555 | 0.960 |
| Medial occipital | 13,239 | -0.004 | 0.018 | -0.230 | 0.817 | 0.960 |
| ***Subcortical volumes*** | |  |  |  |  |  |
| Thalamus | 13,224 | 0.004 | 0.014 | 0.320 | 0.750 | 0.282 |
| Caudate | 13,212 | 0.032 | 0.018 | 1.800 | 0.072 | 0.989 |
| Putamen | 13,210 | 0.032 | 0.017 | 1.930 | 0.054 | 0.846 |
| Pallidum | 13,233 | 0.015 | 0.016 | 0.970 | 0.333 | 0.559 |
| Hippocampus | 13,235 | -0.010 | 0.016 | -0.610 | 0.543 | 0.778 |
| Amygdala | 13,235 | -0.031 | 0.015 | -2.010 | 0.044 | 0.088 |
| Accumbens | 13,237 | -0.001 | 0.016 | -0.080 | 0.938 | 0.559 |
| left hippocampus | 13,203 | -0.008 | 0.017 | -0.480 | 0.631 | 0.380 |
| right hippocampus | 13,200 | -0.010 | 0.017 | -0.590 | 0.557 | 0.989 |
| left accumbens | 13,193 | 0.007 | 0.018 | 0.390 | 0.699 | 0.989 |
| right accumbens | 13,193 | -0.008 | 0.018 | -0.450 | 0.651 | 0.282 |

#### Table S12. Main effect of birth weight on brain structures in the interaction model.

| **Brain structure** | **N** | **β** | **SE** | **z** | **p_uncorrected_** | **p_corrected_** |
| --- | --- | --- | --- | --- | --- | --- |
| ***Global*** |  |  |  |  |  |  |
| Cortical thickness | 11,889 | -0.026 | 0.013 | -1.970 | 0.049 | 0.049 |
| Surface area | 11,896 | 0.044 | 0.007 | 5.950 | <0.001 | <0.001 |
| Cortical volume | 11,907 | 0.029 | 0.008 | 3.610 | <0.001 | <0.001 |
| ***Lobes*** |  |  |  |  |  |  |
| ***Cortical thickness*** | |  |  |  |  |  |
| Frontal lobe | 11,891 | -0.018 | 0.013 | -1.370 | 0.169 | 0.237 |
| Temporal lobe | 11,908 | -0.053 | 0.013 | -4.140 | <0.001 | <0.001 |
| Occipital lobe | 11,914 | -0.088 | 0.014 | -6.480 | <0.001 | <0.001 |
| Parietal lobe | 11,871 | -0.030 | 0.013 | -2.240 | 0.025 | 0.050 |
| Cingulate lobe | 11,916 | -0.016 | 0.012 | -1.350 | 0.178 | 0.237 |
| Insula lobe | 11,920 | 0.033 | 0.013 | 2.620 | 0.009 | 0.024 |
| Postcentral lobe | 11,918 | -0.009 | 0.013 | -0.670 | 0.502 | 0.574 |
| Paracentral lobe | 11,912 | -0.001 | 0.013 | -0.100 | 0.924 | 0.924 |
| ***Surface area*** | |  |  |  |  |  |
| Frontal lobe | 11,902 | 0.008 | 0.008 | 0.990 | 0.324 | 0.370 |
| Temporal lobe | 11,904 | 0.038 | 0.008 | 4.580 | <0.001 | <0.001 |
| Occipital lobe | 11,898 | 0.013 | 0.012 | 1.120 | 0.264 | 0.352 |
| Parietal lobe | 11,900 | 0.078 | 0.009 | 8.430 | <0.001 | <0.001 |
| Cingulate lobe | 11,912 | 0.060 | 0.009 | 6.720 | <0.001 | <0.001 |
| Insula lobe | 11,912 | 0.029 | 0.010 | 3.000 | 0.003 | 0.005 |
| Postcentral lobe | 11,912 | 0.061 | 0.010 | 6.320 | <0.001 | <0.001 |
| Paracentral lobe | 11,912 | 0.001 | 0.010 | 0.130 | 0.900 | 0.900 |
| ***Cortical volume*** | |  |  |  |  |  |
| Frontal lobe | 11,909 | 0.013 | 0.009 | 1.420 | 0.156 | 0.208 |
| Temporal lobe | 11,906 | 0.010 | 0.009 | 1.070 | 0.285 | 0.326 |
| Occipital lobe | 11,906 | -0.024 | 0.011 | -2.080 | 0.037 | 0.059 |
| Parietal lobe | 11,912 | 0.069 | 0.009 | 7.250 | <0.001 | <0.001 |
| Cingulate lobe | 11,912 | 0.060 | 0.010 | 6.180 | <0.001 | <0.001 |
| Insula lobe | 11,896 | 0.045 | 0.010 | 4.360 | <0.001 | <0.001 |
| Postcentral lobe | 11,910 | 0.046 | 0.011 | 4.330 | <0.001 | <0.001 |
| Paracentral lobe | 11,913 | 0.006 | 0.011 | 0.520 | 0.604 | 0.604 |
| ***Parcellations*** |  |  |  |  |  |  |
| ***Cortical thickness*** | |  |  |  |  |  |
| Caudal anterior cingulate | 11,919 | 0.010 | 0.011 | 0.870 | 0.386 | 0.478 |
| Caudal middle frontal | 11,892 | 0.015 | 0.013 | 1.180 | 0.239 | 0.327 |
| Entorhinal | 11,901 | -0.042 | 0.012 | -3.410 | 0.001 | 0.003 |
| Fusiform | 11,913 | -0.022 | 0.013 | -1.730 | 0.084 | 0.128 |
| Inferior parietal | 11,889 | -0.014 | 0.013 | -1.080 | 0.279 | 0.363 |
| Inferior temporal | 11,917 | -0.048 | 0.013 | -3.690 | <0.001 | <0.001 |
| Isthmus cingulate | 11,917 | -0.029 | 0.013 | -2.350 | 0.019 | 0.038 |
| Lateral occipital | 11,910 | -0.108 | 0.013 | -8.060 | <0.001 | <0.001 |
| Lateral orbitofrontal | 11,911 | -0.029 | 0.013 | -2.250 | 0.025 | 0.046 |
| Medial orbitofrontal | 11,911 | -0.051 | 0.013 | -4.010 | <0.001 | <0.001 |
| Middle temporal | 11,918 | 0.038 | 0.013 | 2.990 | 0.003 | 0.009 |
| Parahippocampal | 11,916 | -0.050 | 0.013 | -3.870 | <0.001 | <0.001 |
| Paracentral | 11,912 | -0.001 | 0.013 | -0.100 | 0.924 | 0.924 |
| Postcentral | 11,918 | -0.009 | 0.013 | -0.670 | 0.502 | 0.593 |
| Posterior cingulate | 11,920 | -0.022 | 0.012 | -1.750 | 0.081 | 0.128 |
| Precentral | 11,899 | 0.002 | 0.013 | 0.130 | 0.898 | 0.924 |
| Precuneus | 11,896 | -0.065 | 0.013 | -4.800 | <0.001 | <0.001 |
| Rostral anterior cingulate | 11,917 | -0.033 | 0.012 | -2.830 | 0.005 | 0.013 |
| Superior parietal | 11,891 | -0.057 | 0.013 | -4.260 | <0.001 | <0.001 |
| Supramarginal | 11,887 | 0.023 | 0.013 | 1.750 | 0.080 | 0.128 |
| Frontal pole | 11,914 | -0.028 | 0.012 | -2.360 | 0.018 | 0.038 |
| Insula | 11,920 | 0.033 | 0.013 | 2.620 | 0.009 | 0.021 |
| Superior temporal | 11,906 | -0.016 | 0.013 | -1.220 | 0.221 | 0.319 |
| Inferior frontal | 11,891 | 0.004 | 0.013 | 0.290 | 0.775 | 0.840 |
| Dorsal lateral prefrontal | 11,875 | 0.006 | 0.013 | 0.480 | 0.632 | 0.714 |
| Medial occipital | 11,913 | -0.054 | 0.013 | -4.000 | <0.001 | <0.001 |
| ***Surface area*** | |  |  |  |  |  |
| Caudal anterior cingulate | 11,914 | 0.019 | 0.010 | 1.940 | 0.052 | 0.071 |
| Caudal middle frontal | 11,910 | 0.005 | 0.011 | 0.470 | 0.640 | 0.693 |
| Entorhinal | 11,918 | -0.020 | 0.012 | -1.680 | 0.092 | 0.120 |
| Fusiform | 11,911 | 0.026 | 0.010 | 2.720 | 0.006 | 0.010 |
| Inferior parietal | 11,912 | 0.092 | 0.010 | 8.760 | <0.001 | <0.001 |
| Inferior temporal | 11,912 | 0.051 | 0.010 | 5.210 | <0.001 | <0.001 |
| Isthmus cingulate | 11,910 | 0.055 | 0.010 | 5.500 | <0.001 | <0.001 |
| Lateral occipital | 11,907 | 0.008 | 0.011 | 0.770 | 0.439 | 0.496 |
| Lateral orbitofrontal | 11,911 | 0.059 | 0.010 | 5.940 | <0.001 | <0.001 |
| Medial orbitofrontal | 11,912 | 0.051 | 0.009 | 5.780 | <0.001 | <0.001 |
| Middle temporal | 11,904 | 0.097 | 0.010 | 9.930 | <0.001 | <0.001 |
| Parahippocampal | 11,913 | 0.017 | 0.011 | 1.610 | 0.108 | 0.134 |
| Paracentral | 11,912 | 0.001 | 0.010 | 0.130 | 0.900 | 0.900 |
| Postcentral | 11,912 | 0.061 | 0.010 | 6.320 | <0.001 | <0.001 |
| Posterior cingulate | 11,910 | 0.051 | 0.010 | 5.230 | <0.001 | <0.001 |
| Precentral | 11,906 | 0.002 | 0.010 | 0.230 | 0.815 | 0.848 |
| Precuneus | 11,901 | 0.035 | 0.010 | 3.440 | 0.001 | 0.002 |
| Rostral anterior cingulate | 11,913 | 0.057 | 0.010 | 5.750 | <0.001 | <0.001 |
| Superior parietal | 11,904 | 0.057 | 0.011 | 5.260 | <0.001 | <0.001 |
| Supramarginal | 11,910 | 0.062 | 0.010 | 6.220 | <0.001 | <0.001 |
| Frontal pole | 11,916 | 0.058 | 0.010 | 5.650 | <0.001 | <0.001 |
| Insula | 11,912 | 0.029 | 0.010 | 3.000 | 0.003 | 0.005 |
| Superior temporal | 11,910 | 0.025 | 0.009 | 2.590 | 0.009 | 0.014 |
| Inferior frontal | 11,910 | 0.034 | 0.011 | 3.190 | 0.001 | 0.002 |
| Dorsal lateral prefrontal | 11,900 | -0.018 | 0.009 | -2.030 | 0.042 | 0.061 |
| Medial occipital | 11,907 | 0.013 | 0.013 | 1.040 | 0.298 | 0.352 |
| ***Cortical volume*** | |  |  |  |  |  |
| Caudal anterior cingulate | 11,918 | 0.015 | 0.010 | 1.400 | 0.161 | 0.220 |
| Caudal middle frontal | 11,912 | 0.012 | 0.011 | 1.130 | 0.260 | 0.319 |
| Entorhinal | 11,912 | -0.028 | 0.012 | -2.340 | 0.019 | 0.031 |
| Fusiform | 11,914 | 0.014 | 0.010 | 1.320 | 0.186 | 0.242 |
| Inferior parietal | 11,914 | 0.093 | 0.011 | 8.830 | <0.001 | <0.001 |
| Inferior temporal | 11,914 | 0.026 | 0.010 | 2.630 | 0.009 | 0.018 |
| Isthmus cingulate | 11,915 | 0.055 | 0.010 | 5.230 | <0.001 | <0.001 |
| Lateral occipital | 11,912 | -0.040 | 0.011 | -3.760 | <0.001 | <0.001 |
| Lateral orbitofrontal | 11,905 | 0.044 | 0.010 | 4.250 | <0.001 | <0.001 |
| Medial orbitofrontal | 11,916 | 0.025 | 0.010 | 2.490 | 0.013 | 0.024 |
| Middle temporal | 11,910 | 0.110 | 0.010 | 11.080 | <0.001 | <0.001 |
| Parahippocampal | 11,916 | -0.035 | 0.012 | -2.830 | 0.005 | 0.011 |
| Paracentral | 11,913 | 0.006 | 0.011 | 0.520 | 0.604 | 0.654 |
| Postcentral | 11,910 | 0.046 | 0.011 | 4.330 | <0.001 | <0.001 |
| Posterior cingulate | 11,916 | 0.054 | 0.010 | 5.150 | <0.001 | <0.001 |
| Precentral | 11,912 | 0.010 | 0.011 | 0.910 | 0.361 | 0.408 |
| Precuneus | 11,903 | 0.015 | 0.010 | 1.500 | 0.134 | 0.194 |
| Rostral anterior cingulate | 11,917 | 0.057 | 0.010 | 5.450 | <0.001 | <0.001 |
| Superior parietal | 11,915 | 0.027 | 0.011 | 2.430 | 0.015 | 0.026 |
| Supramarginal | 11,911 | 0.073 | 0.010 | 7.160 | <0.001 | <0.001 |
| Frontal pole | 11,906 | 0.021 | 0.011 | 1.860 | 0.063 | 0.096 |
| Insula | 11,896 | 0.045 | 0.010 | 4.360 | <0.001 | <0.001 |
| Superior temporal | 11,901 | -0.001 | 0.010 | -0.130 | 0.895 | 0.895 |
| Inferior frontal | 11,906 | 0.042 | 0.011 | 3.870 | <0.001 | <0.001 |
| Dorsal lateral prefrontal | 11,909 | -0.010 | 0.009 | -1.100 | 0.270 | 0.319 |
| Medial occipital | 11,904 | 0.003 | 0.012 | 0.250 | 0.800 | 0.832 |
| ***Subcortical volumes*** | |  |  |  |  |  |
| Thalamus | 11,894 | 0.049 | 0.010 | 5.130 | <0.001 | <0.001 |
| Caudate | 11,890 | 0.002 | 0.012 | 0.200 | 0.845 | 0.845 |
| Putamen | 11,881 | -0.013 | 0.012 | -1.170 | 0.243 | 0.297 |
| Pallidum | 11,904 | 0.053 | 0.011 | 4.800 | <0.001 | <0.001 |
| Hippocampus | 11,908 | 0.015 | 0.011 | 1.380 | 0.167 | 0.230 |
| Amygdala | 11,902 | -0.032 | 0.011 | -3.090 | 0.002 | 0.004 |
| Accumbens | 11,911 | 0.102 | 0.011 | 9.000 | <0.001 | <0.001 |
| left hippocampus | 11,877 | 0.010 | 0.012 | 0.810 | 0.418 | 0.460 |
| right hippocampus | 11,879 | 0.021 | 0.012 | 1.780 | 0.075 | 0.118 |
| left accumbens | 11,873 | 0.102 | 0.012 | 8.170 | <0.001 | <0.001 |
| right accumbens | 11,872 | 0.099 | 0.013 | 7.860 | <0.001 | <0.001 |

#### Table S13. Main effect of Townsend deprivation index on brain structures in the interaction model.

| **Brain structure** | **N** | **β** | **SE** | **z** | **p_uncorrected_** | **p_corrected_** |
| --- | --- | --- | --- | --- | --- | --- |
| ***Global*** |  |  |  |  |  |  |
| Cortical thickness | 18,016 | -0.007 | 0.003 | -2.620 | 0.009 | 0.020 |
| Surface area | 18,037 | -0.001 | 0.001 | -0.660 | 0.510 | 0.510 |
| Cortical volume | 18,050 | -0.004 | 0.002 | -2.480 | 0.013 | 0.020 |
| ***Lobes*** |  |  |  |  |  |  |
| ***Cortical thickness*** | |  |  |  |  |  |
| Frontal lobe | 18,019 | -0.006 | 0.003 | -2.570 | 0.010 | 0.027 |
| Temporal lobe | 18,044 | -0.008 | 0.002 | -3.450 | 0.001 | 0.008 |
| Occipital lobe | 18,061 | 0.002 | 0.003 | 0.940 | 0.345 | 0.345 |
| Parietal lobe | 17,989 | -0.007 | 0.003 | -2.960 | 0.003 | 0.012 |
| Cingulate lobe | 18,066 | -0.005 | 0.002 | -2.080 | 0.037 | 0.067 |
| Insula lobe | 18,069 | -0.005 | 0.002 | -2.030 | 0.042 | 0.067 |
| Postcentral lobe | 18,059 | -0.004 | 0.002 | -1.560 | 0.119 | 0.159 |
| Paracentral lobe | 18,058 | -0.003 | 0.003 | -1.010 | 0.311 | 0.345 |
| ***Surface area*** |  |  |  |  |  |  |
| Frontal lobe | 18,042 | -0.001 | 0.002 | -0.530 | 0.597 | 0.796 |
| Temporal lobe | 18,047 | -0.001 | 0.002 | -0.710 | 0.478 | 0.796 |
| Occipital lobe | 18,043 | -0.002 | 0.002 | -0.920 | 0.356 | 0.796 |
| Parietal lobe | 18,044 | -0.001 | 0.002 | -0.340 | 0.734 | 0.839 |
| Cingulate lobe | 18,058 | 0.003 | 0.002 | 1.620 | 0.104 | 0.796 |
| Insula lobe | 18,057 | 0.002 | 0.002 | 0.930 | 0.350 | 0.796 |
| Postcentral lobe | 18,057 | <0.001 | 0.002 | -0.020 | 0.984 | 0.984 |
| Paracentral lobe | 18,055 | 0.001 | 0.002 | 0.540 | 0.586 | 0.796 |
| ***Cortical volume*** |  |  |  |  |  |  |
| Frontal lobe | 18,048 | -0.005 | 0.002 | -2.690 | 0.007 | 0.028 |
| Temporal lobe | 18,052 | -0.004 | 0.002 | -2.320 | 0.020 | 0.050 |
| Occipital lobe | 18,053 | 0.001 | 0.002 | 0.240 | 0.812 | 0.928 |
| Parietal lobe | 18,058 | -0.005 | 0.002 | -2.940 | 0.003 | 0.024 |
| Cingulate lobe | 18,060 | <0.001 | 0.002 | 0.030 | 0.979 | 0.979 |
| Insula lobe | 18,038 | -0.004 | 0.002 | -2.250 | 0.025 | 0.050 |
| Postcentral lobe | 18,056 | -0.003 | 0.002 | -1.370 | 0.172 | 0.275 |
| Paracentral lobe | 18,062 | -0.001 | 0.002 | -0.250 | 0.804 | 0.928 |
| ***Parcellations*** |  |  |  |  |  |  |
| ***Cortical thickness*** | |  |  |  |  |  |
| Caudal anterior cingulate | 18,069 | -0.002 | 0.002 | -0.790 | 0.428 | 0.464 |
| Caudal middle frontal | 18,022 | -0.006 | 0.002 | -2.610 | 0.009 | 0.021 |
| Entorhinal | 18,036 | -0.006 | 0.002 | -2.480 | 0.013 | 0.026 |
| Fusiform | 18,054 | -0.008 | 0.002 | -3.150 | 0.002 | 0.007 |
| Inferior parietal | 18,015 | -0.008 | 0.002 | -3.320 | 0.001 | 0.005 |
| Inferior temporal | 18,062 | -0.006 | 0.002 | -2.450 | 0.014 | 0.026 |
| Isthmus cingulate | 18,066 | -0.003 | 0.002 | -1.250 | 0.211 | 0.249 |
| Lateral occipital | 18,058 | 0.001 | 0.003 | 0.300 | 0.764 | 0.780 |
| Lateral orbitofrontal | 18,055 | -0.008 | 0.002 | -3.260 | 0.001 | 0.005 |
| Medial orbitofrontal | 18,057 | -0.008 | 0.002 | -3.430 | 0.001 | 0.005 |
| Middle temporal | 18,066 | -0.007 | 0.002 | -2.770 | 0.006 | 0.020 |
| Parahippocampal | 18,062 | -0.006 | 0.002 | -2.630 | 0.009 | 0.021 |
| Paracentral | 18,058 | -0.003 | 0.003 | -1.010 | 0.311 | 0.352 |
| Postcentral | 18,059 | -0.004 | 0.002 | -1.560 | 0.119 | 0.147 |
| Posterior cingulate | 18,068 | -0.006 | 0.002 | -2.710 | 0.007 | 0.020 |
| Precentral | 18,036 | -0.008 | 0.002 | -3.140 | 0.002 | 0.007 |
| Precuneus | 18,031 | -0.006 | 0.003 | -2.270 | 0.023 | 0.040 |
| Rostral anterior cingulate | 18,065 | -0.004 | 0.002 | -1.670 | 0.095 | 0.124 |
| Superior parietal | 18,017 | -0.005 | 0.003 | -1.870 | 0.061 | 0.083 |
| Supramarginal | 18,015 | -0.010 | 0.002 | -4.180 | <0.001 | <0.001 |
| Frontal pole | 18,058 | -0.001 | 0.002 | -0.280 | 0.780 | 0.780 |
| Insula | 18,069 | -0.005 | 0.002 | -2.030 | 0.042 | 0.061 |
| Superior temporal | 18,041 | -0.005 | 0.003 | -2.170 | 0.030 | 0.049 |
| Inferior frontal | 18,021 | -0.008 | 0.002 | -3.380 | 0.001 | 0.005 |
| Dorsal lateral prefrontal | 17,984 | -0.006 | 0.002 | -2.460 | 0.014 | 0.026 |
| Medial occipital | 18,059 | 0.005 | 0.003 | 2.080 | 0.037 | 0.057 |
| ***Surface area*** |  |  |  |  |  |  |
| Caudal anterior cingulate | 18,061 | 0.002 | 0.002 | 1.040 | 0.296 | 0.758 |
| Caudal middle frontal | 18,051 | <0.001 | 0.002 | 0.180 | 0.861 | 0.940 |
| Entorhinal | 18,065 | -0.001 | 0.002 | -0.560 | 0.577 | 0.922 |
| Fusiform | 18,059 | -0.002 | 0.002 | -1.130 | 0.258 | 0.745 |
| Inferior parietal | 18,062 | -0.003 | 0.002 | -1.560 | 0.119 | 0.650 |
| Inferior temporal | 18,061 | -0.004 | 0.002 | -2.220 | 0.026 | 0.338 |
| Isthmus cingulate | 18,052 | 0.001 | 0.002 | 0.310 | 0.757 | 0.932 |
| Lateral occipital | 18,054 | -0.001 | 0.002 | -0.510 | 0.610 | 0.922 |
| Lateral orbitofrontal | 18,059 | -0.004 | 0.002 | -2.250 | 0.024 | 0.338 |
| Medial orbitofrontal | 18,058 | -0.001 | 0.002 | -0.410 | 0.685 | 0.932 |
| Middle temporal | 18,051 | -0.003 | 0.002 | -1.840 | 0.066 | 0.572 |
| Parahippocampal | 18,060 | -0.002 | 0.002 | -0.830 | 0.406 | 0.812 |
| Paracentral | 18,055 | 0.001 | 0.002 | 0.540 | 0.586 | 0.922 |
| Postcentral | 18,057 | <0.001 | 0.002 | -0.020 | 0.984 | 0.984 |
| Posterior cingulate | 18,053 | 0.002 | 0.002 | 1.190 | 0.235 | 0.745 |
| Precentral | 18,055 | -0.002 | 0.002 | -1.310 | 0.190 | 0.706 |
| Precuneus | 18,040 | 0.001 | 0.002 | 0.340 | 0.732 | 0.932 |
| Rostral anterior cingulate | 18,060 | 0.003 | 0.002 | 1.540 | 0.125 | 0.650 |
| Superior parietal | 18,049 | <0.001 | 0.002 | 0.170 | 0.868 | 0.940 |
| Supramarginal | 18,056 | 0.001 | 0.002 | 0.270 | 0.789 | 0.932 |
| Frontal pole | 18,067 | -0.001 | 0.002 | -0.760 | 0.450 | 0.836 |
| Insula | 18,057 | 0.002 | 0.002 | 0.930 | 0.350 | 0.758 |
| Superior temporal | 18,057 | 0.002 | 0.002 | 1.330 | 0.184 | 0.706 |
| Inferior frontal | 18,050 | 0.001 | 0.002 | 0.470 | 0.638 | 0.922 |
| Dorsal lateral prefrontal | 18,041 | <0.001 | 0.002 | -0.050 | 0.962 | 0.984 |
| Medial occipital | 18,050 | -0.002 | 0.002 | -0.950 | 0.342 | 0.758 |
| ***Cortical volume*** |  |  |  |  |  |  |
| Caudal anterior cingulate | 18,069 | <0.001 | 0.002 | 0.240 | 0.807 | 0.906 |
| Caudal middle frontal | 18,055 | -0.003 | 0.002 | -1.300 | 0.195 | 0.338 |
| Entorhinal | 18,058 | <0.001 | 0.002 | -0.220 | 0.829 | 0.906 |
| Fusiform | 18,062 | -0.005 | 0.002 | -2.400 | 0.016 | 0.059 |
| Inferior parietal | 18,061 | -0.007 | 0.002 | -3.560 | <0.001 | <0.001 |
| Inferior temporal | 18,062 | -0.005 | 0.002 | -2.480 | 0.013 | 0.056 |
| Isthmus cingulate | 18,062 | -0.001 | 0.002 | -0.310 | 0.760 | 0.906 |
| Lateral occipital | 18,058 | <0.001 | 0.002 | -0.210 | 0.836 | 0.906 |
| Lateral orbitofrontal | 18,047 | -0.007 | 0.002 | -3.540 | <0.001 | <0.001 |
| Medial orbitofrontal | 18,064 | -0.006 | 0.002 | -2.960 | 0.003 | 0.016 |
| Middle temporal | 18,056 | -0.006 | 0.002 | -3.130 | 0.002 | 0.013 |
| Parahippocampal | 18,065 | -0.005 | 0.002 | -2.130 | 0.033 | 0.095 |
| Paracentral | 18,062 | -0.001 | 0.002 | -0.250 | 0.804 | 0.906 |
| Postcentral | 18,056 | -0.003 | 0.002 | -1.370 | 0.172 | 0.319 |
| Posterior cingulate | 18,063 | -0.001 | 0.002 | -0.590 | 0.552 | 0.844 |
| Precentral | 18,056 | -0.008 | 0.002 | -3.990 | <0.001 | <0.001 |
| Precuneus | 18,038 | -0.003 | 0.002 | -1.400 | 0.162 | 0.319 |
| Rostral anterior cingulate | 18,067 | <0.001 | 0.002 | 0.050 | 0.960 | 0.960 |
| Superior parietal | 18,062 | -0.003 | 0.002 | -1.460 | 0.145 | 0.314 |
| Supramarginal | 18,057 | -0.004 | 0.002 | -2.000 | 0.045 | 0.117 |
| Frontal pole | 18,044 | <0.001 | 0.002 | -0.050 | 0.958 | 0.960 |
| Insula | 18,038 | -0.004 | 0.002 | -2.250 | 0.025 | 0.081 |
| Superior temporal | 18,045 | -0.001 | 0.002 | -0.410 | 0.681 | 0.906 |
| Inferior frontal | 18,051 | -0.002 | 0.002 | -1.150 | 0.251 | 0.408 |
| Dorsal lateral prefrontal | 18,051 | -0.003 | 0.002 | -1.460 | 0.143 | 0.314 |
| Medial occipital | 18,049 | 0.001 | 0.002 | 0.440 | 0.660 | 0.906 |
| ***Subcortical volumes*** | |  |  |  |  |  |
| Thalamus | 18,037 | -0.005 | 0.002 | -2.510 | 0.012 | 0.059 |
| Caudate | 18,009 | <0.001 | 0.002 | 0.050 | 0.963 | 0.963 |
| Putamen | 18,012 | 0.002 | 0.002 | 0.690 | 0.492 | 0.541 |
| Pallidum | 18,044 | -0.005 | 0.002 | -2.570 | 0.010 | 0.059 |
| Hippocampus | 18,047 | -0.004 | 0.002 | -1.990 | 0.046 | 0.084 |
| Amygdala | 18,049 | -0.004 | 0.002 | -1.860 | 0.063 | 0.099 |
| Accumbens | 18,049 | -0.005 | 0.002 | -2.160 | 0.031 | 0.068 |
| left hippocampus | 17,997 | -0.005 | 0.002 | -2.180 | 0.029 | 0.068 |
| right hippocampus | 17,995 | -0.004 | 0.002 | -1.710 | 0.088 | 0.121 |
| left accumbens | 17,985 | -0.004 | 0.002 | -1.490 | 0.137 | 0.167 |
| right accumbens | 17,988 | -0.006 | 0.002 | -2.410 | 0.016 | 0.059 |
